# Evaluating Deep Learning Sepsis Prediction Models in ICUs Under Distribution Shift: A Multi-Centre Retrospective Cohort Study

**DOI:** 10.1101/2025.07.31.25332542

**Authors:** Fanny Tranchellini, Youssef Farag, Catherine Jutzeler, Lakmal Meegahapola

**Affiliations:** Department of Health Sciences and Technology, ETH Zurich, Zurich, Switzerland; Swiss Institute of Bioinformatics, Lausanne, Switzerland

## Abstract

Sepsis remains a leading cause of mortality in intensive care units (ICUs) worldwide, underscoring the urgent need for early detection to improve patient outcomes. While artificial intelligence (AI) models trained on ICU data show promise for sepsis prediction, their clinical utility is frequently hampered by poor generalization under external validation, largely attributable to distribution shifts arising from heterogeneity in data. Prior studies have focused on direct model deployment or conventional transfer learning methods (e.g., fine-tuning), yet systematic exploration of alternative strategies and root causes of performance degradation remains limited. In this study, we quantify those distribution shifts across three harmonized adult ICU cohorts: the high-resolution HiRID database (Bern University Hospital, Switzerland; 29 698 stays, 2008–2019; 6.3 % sepsis), MIMIC-IV (Beth Israel Deaconess Medical Center, USA; 63 425 stays, 2008–2019; 5.2 % sepsis), and eICU (208 US hospitals; 123 413 stays, 2014–2015; 4.6 % sepsis) for a total of 216 536 stays and 10 846 sepsis cases. We then evaluate five deployment strategies across three model architectures (CNN, InceptionTime, LSTM) under four target-data regimes: *none*, *small* (*<* 8000 stays), *medium* (8000–32000), and *large* (*>* 32000). The strategies are direct generalisation, standard transfer learning (fine-tuning / retraining), target training, supervised domain adaptation (DA: MMD or CORAL), and fusion training (merged datasets). Key results demonstrate that fine-tuning consistently underperforms across all data sizes (adjusted p < 0.05 vs. DA, fusion, and retraining) even though it has been the go to method in many prior studies that explored this direction. Retraining and fusion training excel in small and large target domains, while supervised DA methods dominate in medium-sized datasets. For example, DA with maximum mean discrepancy (DA MMD) achieves superior performance in both area under the receiver operating characteristic curve (AUROC = 0.720) and normalized area under the precision-recall curve (nAUPRC = 2.352) compared to fusion training (AUROC = 0.712, nAUPRC = 2.215; p = 0.02, adjusted p = 0.07). Retraining remains competitive (AUROC = 0.719, nAUPRC = 2.326; p > 0.05 vs. DA MMD) but lags in nAUPRC. Overall, our results call for moving beyond routine fine-tuning: retraining or fusion are preferable in data-poor or data-rich scenarios, whereas domain adaptation offers the most stable and substantial gains when moderate target data are available.

## 1 Introduction

Sepsis is a life-threatening disease and one of the leading causes of hospital mortality worldwide, particularly within intensive care units (ICU)^1–4^. It occurs when an infection triggers a dysregulated immune response, leading to widespread inflammation, tissue damage, and impaired organ function^4^. Sepsis is difficult to diagnose early due to its rapid progression and varied clinical presentation; however, timely detection is critical to improve patient outcomes^2–5^. Artificial intelligence (AI), particularly deep learning (DL), offers a promising approach to early detection of sepsis by identifying predictive patterns within the extensive clinical data generated in ICUs, including vital signs, laboratory results, and demographics of patients^3,6–9^. However, a key barrier to the widespread clinical adoption of AI-based predictive models is their poor generalizability between sites. While models typically perform well at their training sites, their accuracy tends to decline at external sites due to *distribution shifts*^10^. These shifts are due to variations in data characteristics, such as differences in vital sign distributions, labeling practices, patient demographics, and clinical protocols, between training (source domain) and deployment (target domain) sites^11^. This challenge was highlighted by a study that showed substantial performance deterioration of a commercial sepsis prediction model, where the model performance dropped to an AUROC of only 0.63 when externally validated in multiple hospitals in the United States, a performance far lower than what the company that developed the model quoted^12^.

Although the need to generalize across sites is widely recognized as a critical challenge, most existing sepsis prediction studies focus narrowly on two simplistic deployment scenarios: external validation and standard transfer learning with fine-tuning. External validation is particularly important when data from the target site are unavailable, and a model trained on source datasets must be deployed without modification. In such cases, the expectation is that the model should demonstrate robustness in different clinical environments. Then, standard transfer learning approaches assume a minor distribution shift between the source and target domains, and therefore only use the small amount of target data to adjust the models to the target setting^13^. Hence, both of these approaches assume that a pre-trained model is available for deployment at the target site. However, other potentially useful strategies, such as supervised domain adaptation (DA), remain largely unexplored when source and target data are available^1,10,14^. This gap is particularly problematic because hospitals differ substantially in their resources, data volumes, data sharing practices, and computational capabilities, which demands flexible strategies suited to specific operational contexts.

For example, real-world scenarios illustrate the importance of tailoring deployment approaches. In a rural hospital setting with limited initial data, a pre-trained model from a larger clinical center may be adopted initially (generalization) and then fine-tuned or fully retrained as local data gradually increase. Ultimately, sufficient local data might enable training of a standalone model from scratch (target training). In a hospital network aiming to standardize sepsis prediction across multiple affiliated institutions, data may be pooled across sites (fusion training), while each site could fine-tune a central model individually or adopt supervised domain adaptation strategies that explicitly address distributional differences between sites. In a nationwide health initiative, an existing model trained in large urban hospitals could be deployed directly (generalization) in smaller rural facilities, with subsequent fine-tuning or retraining once local data accumulate. Such nationwide efforts could also benefit from methods that explicitly account for distributional differences, such as supervised domain adaptation or fusion training, especially when integrating highly diverse datasets. Hence, such different model and data availability across sites based on the operational context (Figure 1**F**), could lead to the need for systematically studying and comparing various deployment strategies (Figure 1**E**): (1) generalization (direct use of a source-trained model without modification), (2) standard transfer learning (fine-tuning: adjusting only selected parameters of the pre-trained model with target data or retraining: updating all model parameters with target data), (3) target training (training with data from the target domain without leveraging any source-domain information), (4) supervised domain adaptation (explicitly aligning feature distributions with alignment techniques such as maximum mean discrepancy (MMD) or correlation alignment (CORAL)), and (5) fusion training (merging source and target datasets into a larger training set); each addressing unique challenges such as data scarcity, domain mismatch, data sharing practices, and practical constraints. Previous studies often neglect these diverse operational contexts in complex clinical environments, rarely perform rigorous evaluations of advanced methods like supervised domain adaptation, or fail to critically compare these strategies in realistic deployment settings^14^.

**Figure 1.**
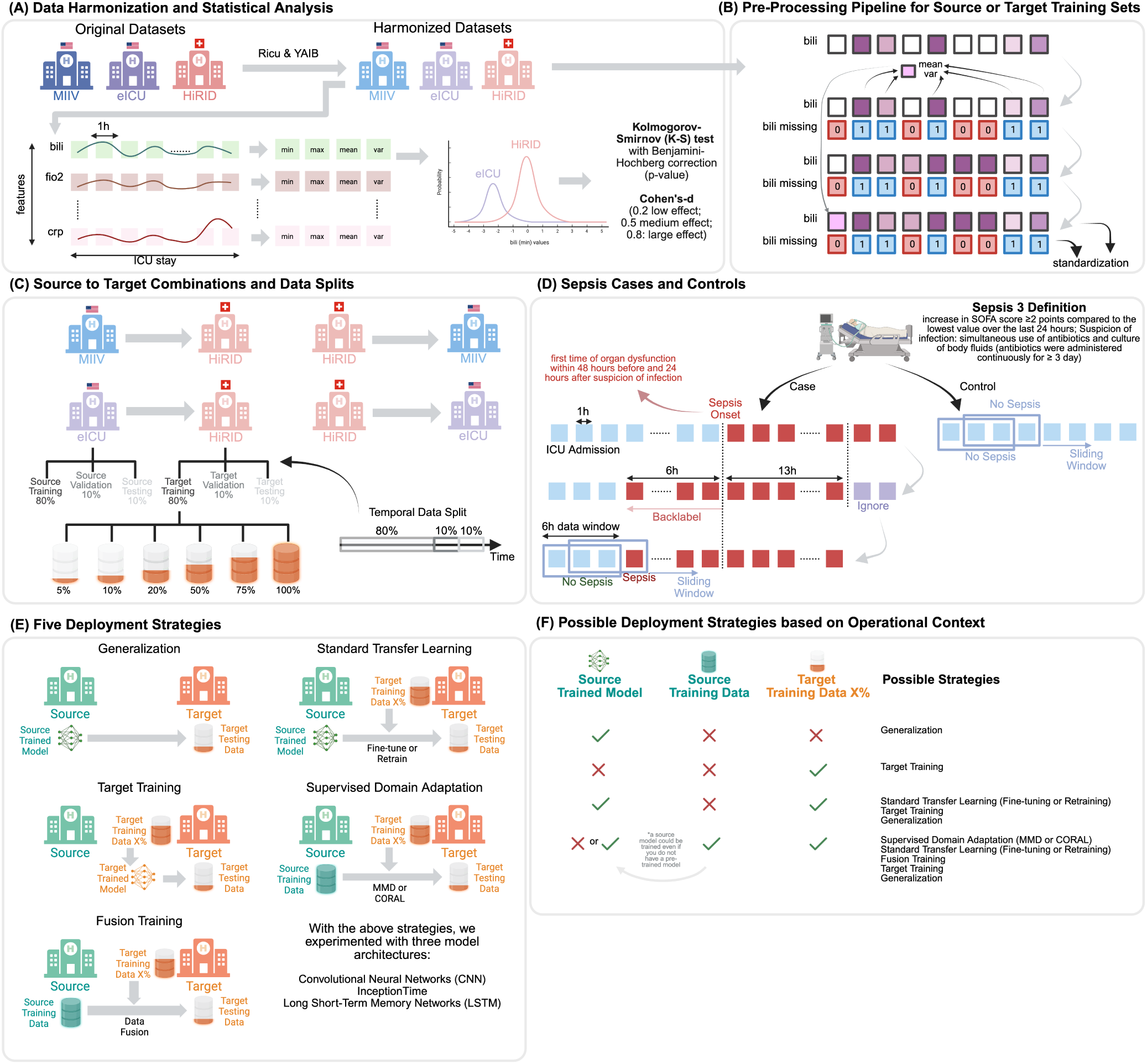
An overview of the main components of the study: **(A)** Original datasets were processed using the RiCU and YAIB pipelines to generate harmonized datasets that allow like-to-like comparisons between the datasets at the feature level. Using these harmonized datasets, we obtained basic statistical properties of patient trajectories, such as minimum, maximum, mean, and variance, at the feature level. These statistical measures were then compared across MIIV-eICU, MIIV-HiRID, and eICU-HiRID combinations using the Kolmogorov-Smirnov (K-S) test and Cohen’s *d*; **(B)** Shows the data pre-processing pipeline used to prepare harmonized data for model training. An example is provided for the feature “bili,” which contains some missing data. Initially, using the available data, the mean and variance are calculated. Then, missing flags are generated as an additional feature column. Next, features are forward-filled, after which any remaining missing data are filled with the mean calculated earlier. Afterwards, all features are standardized. Values learned during this process are applied to both training and validation sets; **(C)** When evaluating model performance, four combinations were considered. For each dataset, we predefined training, validation, and testing datasets. Depending on whether a dataset serves as the source or target in each combination, the relevant training and validation sets were used to train models. Data were split temporally; **(D)** Shows how cases and controls are identified, and how a 6-hour sliding window was used to sample data for model training. We applied a strict version of the Sepsis-3 definition to determine onset inspired from the work by^15^; **(E)** Presents the five main strategies evaluated in this study. For each strategy, it also indicates whether data or models from the source or target domain are utilized; **(F)** Illustrates several realistic operational contexts in terms of model and data availability in the source and target domains for training a model. For each scenario, possible strategies from **(E)** that could be applied are mentioned. This demonstrates that all five broad strategies could be useful in specific setups, making them worth comparing.

In this study, we address these gaps by systematically evaluating the effectiveness of various model deployment strategies for sepsis prediction across heterogeneous ICU settings, with three key contributions: (1) We begin by rigorously quantifying the distribution shifts among three large ICU patient cohorts (MIIV, eICU, and HiRID) to clarify how variations in clinical practices, patient demographics, and data acquisition methods influence their statistical properties, a crucial foundation for understanding generalizability; (2) We then assess cross-domain performance by training on one ICU (source) and validating on another (target) in four pairings: HiRID-MIIV, HiRID-eICU, eICU-HiRID, and MIIV-HiRID; and (3) Finally, to mirror real-world deployments, we analyzed two practical contexts: one in which only a pre-trained source model is available and one that additionally grants access to the source-domain data, and we trace performance as progressively larger fractions of target data become available (5%, 10%, 20%, 50%, 75%, 100%). Under these conditions, we compare five deployment options: direct generalization, fine-tuning, full re-training, supervised domain adaptation (MMD/CORAL), and naive fusion training. Our results show a clear hierarchy that depends on the availability of target data. When data are extremely scarce (*≤*10%) retraining of the source model is the most reliable strategy, with fusion training a close runner-up; both outperform fine-tuning and direct generalization by up to 14% AUROC. At moderate data volumes (*≈* 20–50%) supervised domain adaptation methods become dominant, producing 3–8% AUROC and 0.2–0.6 normalized AUPRC gains over retraining and fine-tuning, while also exhibiting the lowest variance. Once large target datasets (*≥*75%) are available, models trained wholly or partly on the target cohort, that is target training or fusion, match or surpass every transfer learning alternative. Throughout, the benefit of any strategy is modulated by dataset similarity: eICU-trained models generalize better to HiRID than MIIV-trained ones, whereas HiRID models need adaptation before performing well in US cohorts. These findings caution against the default reliance on fine-tuning and highlight the need for deployment plans that adapt to domain shift, data scale, and resource constraints, guidance that could extend beyond sepsis prediction to a broad range of critical care applications.

## 2 Results

The results are organized into three key sections: the existence and significance of distribution shifts between sites, the generalizability of deep learning models across diverse ICU cohorts, and the optimal strategies for deploying these models in target domains.

### 2.1 Existence and Significance of Distribution Shifts Between Sites

Our analysis confirmed that covariate distribution shifts are common and often large between ICU datasets. The Kolmogorov-Smirnov (K-S) tests revealed numerous features with significantly different distributions across the three pairwise dataset comparisons (HiRID vs MIIV, HiRID vs eICU, and MIIV vs eICU). Of the 48 dynamic characteristics examined, between 18 and 29 characteristics (37–60%) showed highly significant differences (p < 0.01) in all four summary statistics when comparing two sites. In other words, each pair of ICU cohorts had dozens of variables that did not follow the same patterns. The HiRID–MIIV pairing had 18 features significantly different in mean/variance/min/max (with an additional three features differing in three of those stats), HiRID–eICU had 19 (plus five with partial differences), and MIIV–eICU showed the most with 29 (plus eight partial). These statistical tests underscore that if one trains a model on the data of an ICU, many input variables will behave differently in the setting of another ICU, violating the assumptions of the model.

To confirm the findings, as another alternative method, Cohen’s d offered a detailed perspective on the extent of the shifts in the feature distribution among the three pairings of data sets. In particular, the pairing of the American dataset (MIIV-eICU) exhibited fewer descriptive statistical characteristics with small, medium, or large effect sizes compared to the two Swiss-American pairings (MIIV-HiRID, eICU-HiRID). Figures 2 and 3 illustrate these differences, highlighting that distribution shifts were generally more pronounced when the data were related to Swiss ICU (HiRID). Within these Swiss-American comparisons, 18 and 16 dynamic features showed medium or large effect sizes for MIIV-HiRID and eICU-HiRID, respectively, whereas the MIIV-eICU pairing contained only seven dynamic features with such effect sizes. Specifically, five dynamic features (band form neutrophils (*bnd*), calcium ionized (*cai*), fraction of inspired oxygen (*fio2*), methemoglobin (*methb*), and urine output) had unique distributions in each dataset. Meanwhile, seven other features (albumin (*alb*), chloride (*cl*), diastolic blood pressure (*dbp*), lymphocytes (*lymph*), mean arterial pressure (*map*), mean corpuscular hemoglobin concentration (*mchc*), and respiratory rate (*resp*)) appeared specific to HiRID, in the sense that MIIV and eICU exhibited more similar distributions relative to each other. These observations collectively suggest that datasets originating from the same country (e.g., MIIV and eICU in the United States) have more comparable data characteristics than datasets from different countries (i.e., HiRID in Switzerland vs. the two American datasets).

**Figure 2.**
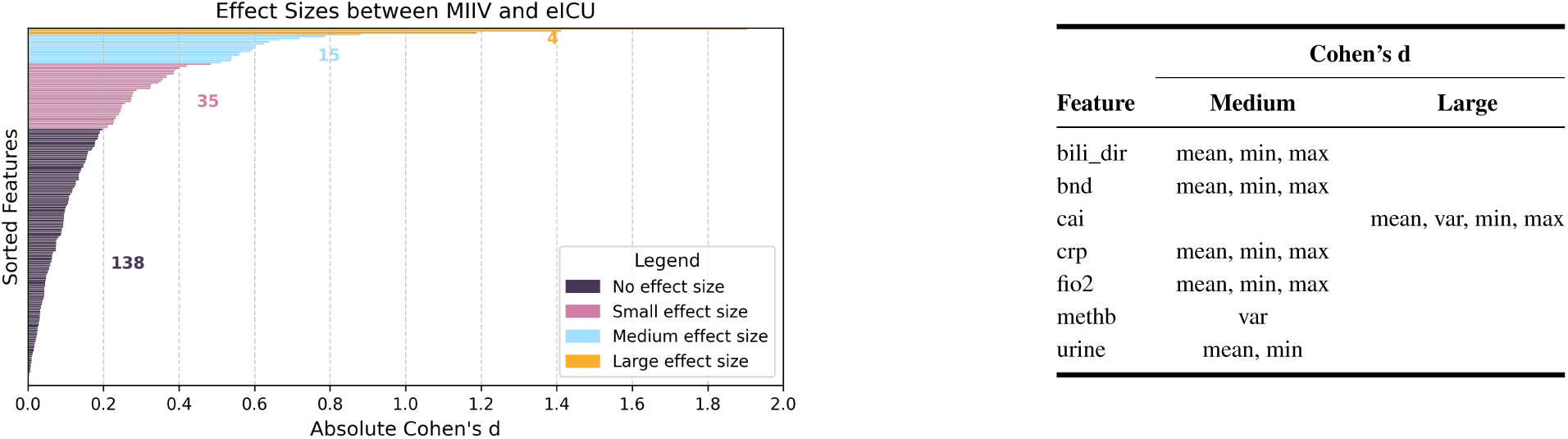
Representation of Cohen’s d analysis of American (MIIV-eICU) dataset pairing. Left: Visualization of the number of features into effect size categories based on absolute Cohen’s d. Right: Classification of features into medium and large effect size categories.

**Figure 3.**
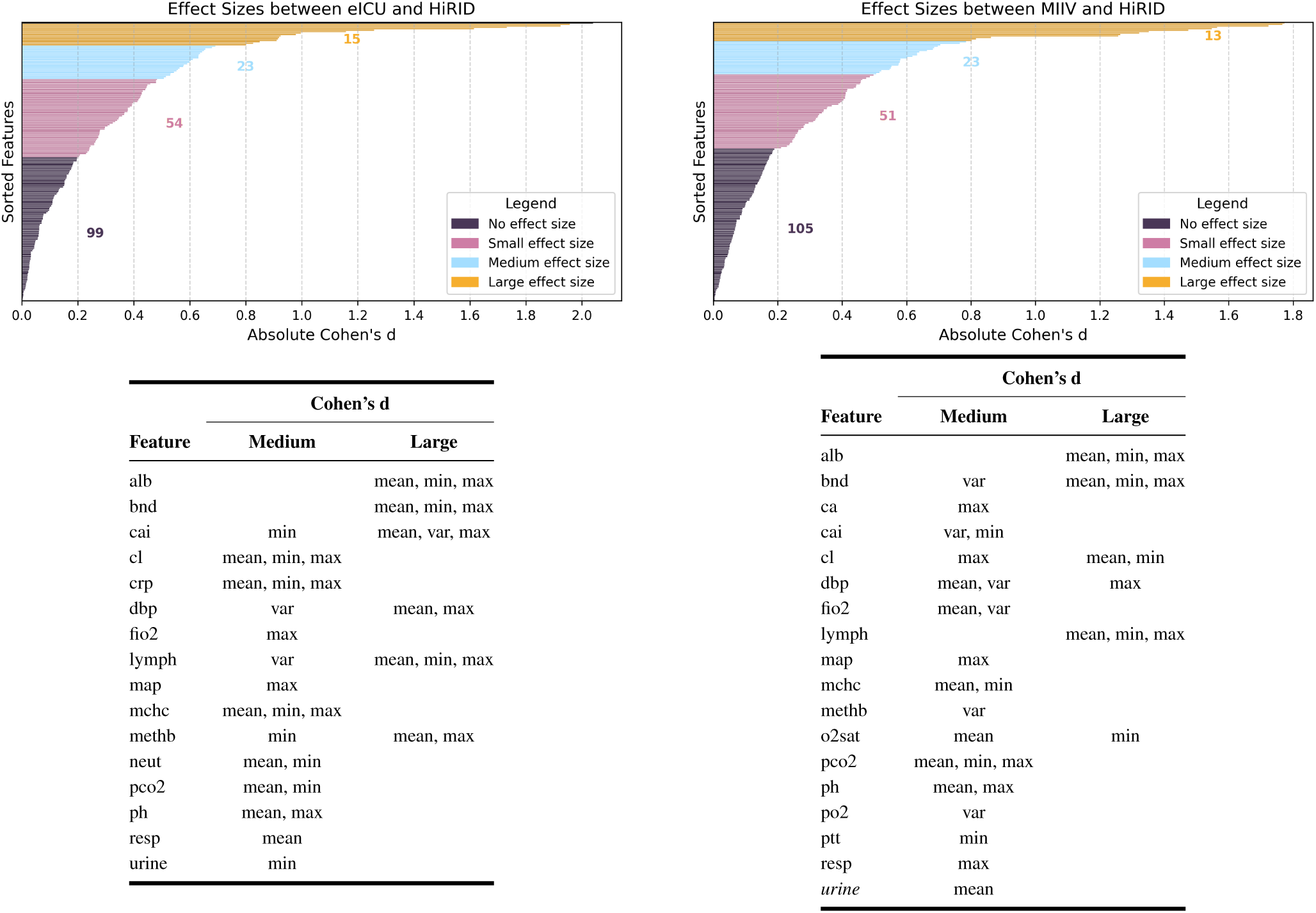
Representation of Cohen’s d analysis of Swiss-American dataset pairings (eICU-HiRID (left) and MIIV-HiRID (right)). Top row: Visualization of the number of features into effect size categories based on absolute Cohen’s d. Bottom row: Classification of features into medium and large effect size categories.

To examine the intricacies of these shifts, we further investigated distribution patterns for several dynamic features, visualizing them through violin plots and stratifying by the outcome of sepsis. For patients who developed sepsis, we also examined measurements before and during the onset of sepsis to see how the distribution might evolve over these time periods. A closer look at these dynamic features through violin plots revealed three key insights about dataset-specific behaviors. First, certain variables showed unique distributions in each dataset (Figure 4): *bnd* (band form neutrophils) and *cai* (ionized calcium) spanned a wider range in HiRID than in MIIV or eICU, while *cai* and *methb* (methemoglobin) were rarely documented in American data sets for patients with sepsis. Furthermore, *fio2* (fraction of inspired oxygen) exhibited multiple distinct peaks whose concentration levels varied between the three cohorts. In Figure 5, other features: *alb* (albumin), *cl* (chloride), *dbp* (diastolic blood pressure), and *mchc* (mean corpuscular hemoglobin concentration), showed a marked shift in the HiRID data set relative to MIIV and eICU, persisting in both sepsis subpopulations. Finally, as shown in Figure 6, *C-reactive protein* displayed a notably wider distribution in the eICU compared to the other two datasets, an observation consistent between the sepsis and non-sepsis groups. These varied distribution patterns highlight the substantial heterogeneity between hospitals that can undermine naive data pooling or model deployment without adaptation, as local definitions, measurement ranges, and clinical protocols for critical laboratory values appear to differ significantly between these ICU settings.

**Figure 4.**
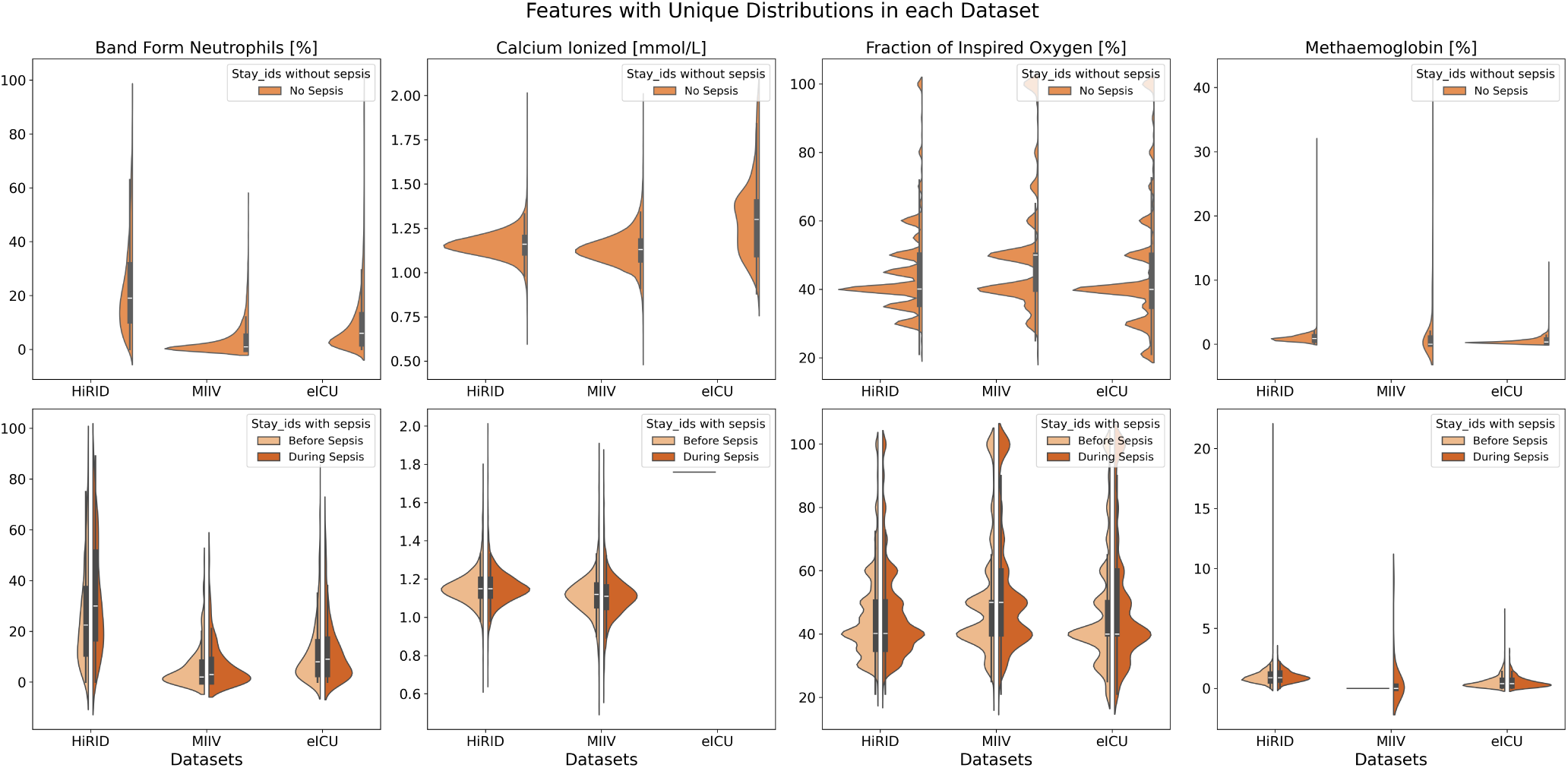
Features with Unique Distributions in each Dataset.

**Figure 5.**
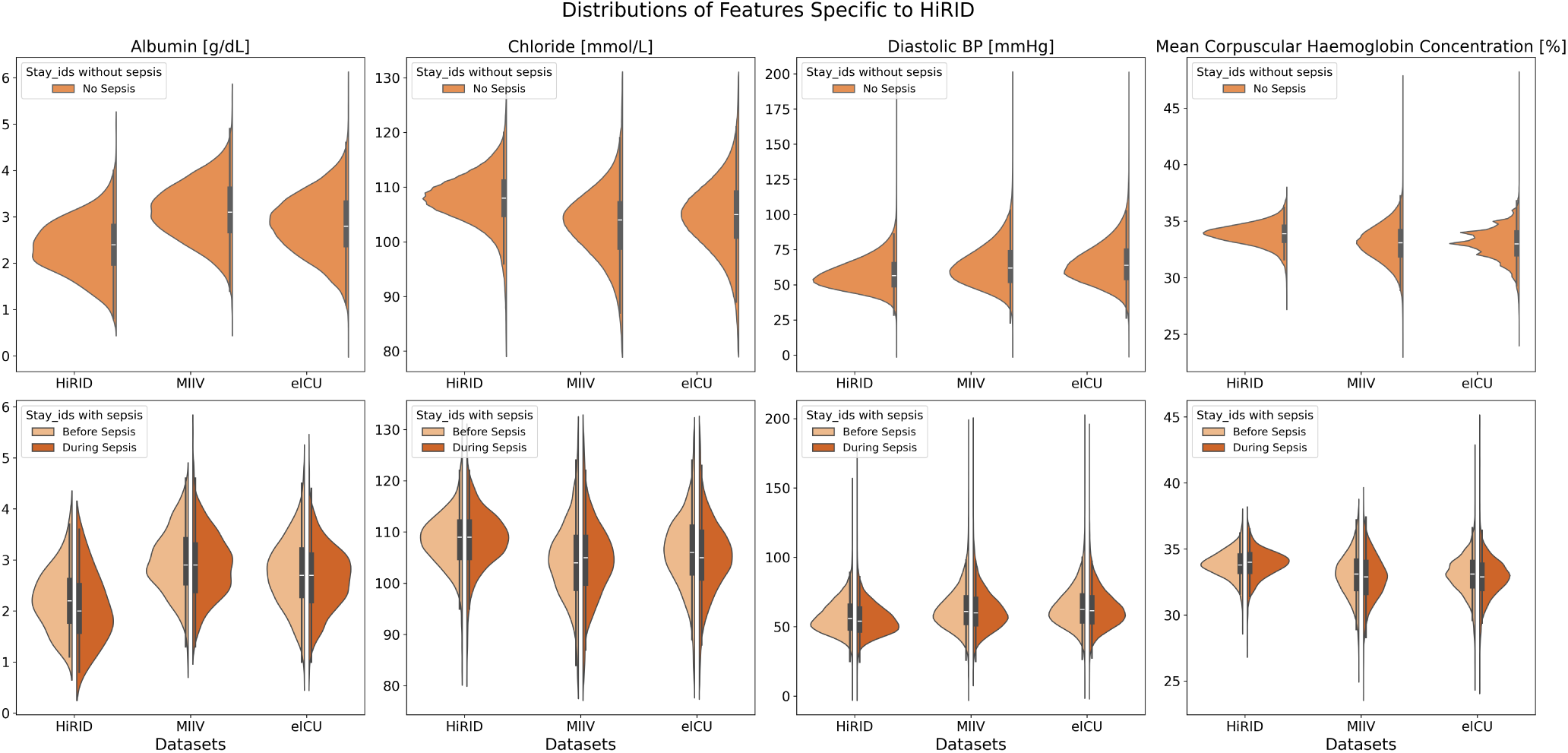
Features with Distributions Specific to HiRID Dataset.

**Figure 6.**
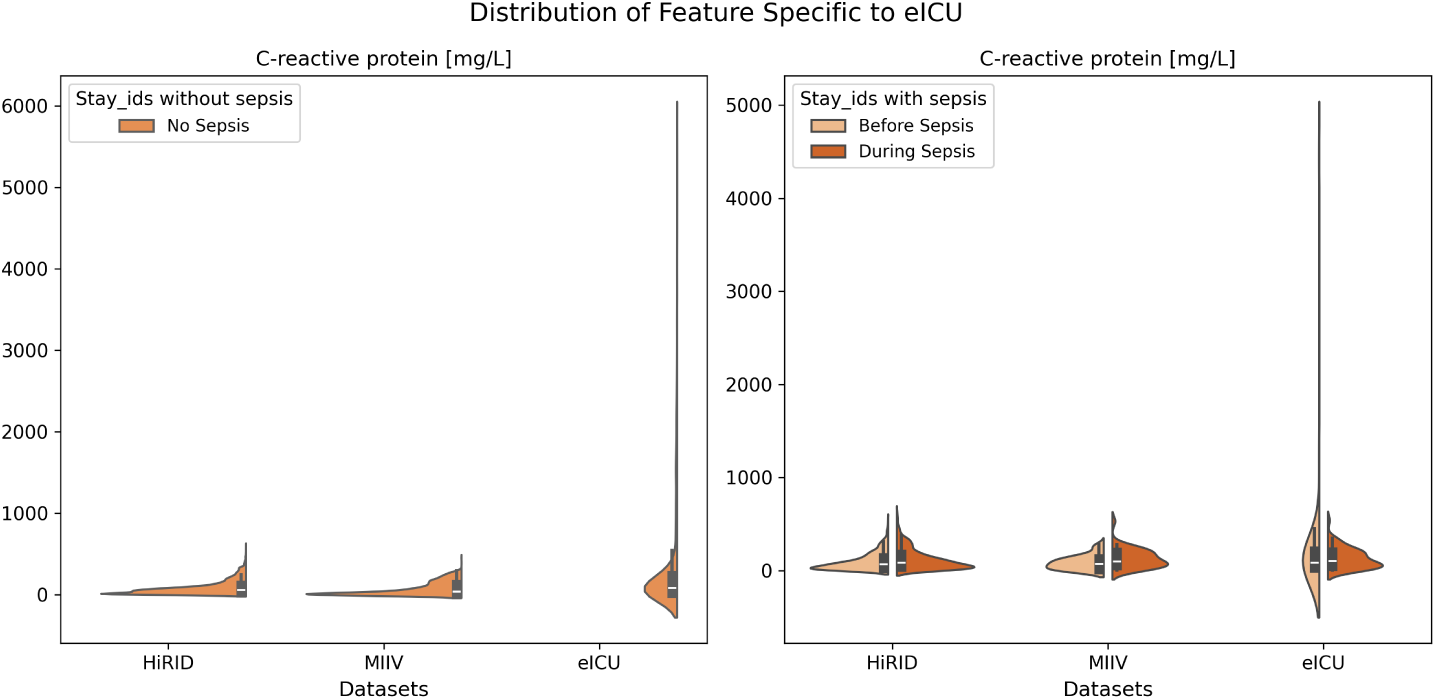
C-Reactive Protein Distribution Specific to eICU Dataset.

Collectively, these findings demonstrate that feature-wise distribution shifts can be large and highly variable in different ICUs, further posing questions of whether naively pooling data (fusion training) or deploying models without adaptation (generalization) may fail to capture subtle, but critical, differences between sites in clinical practice and measurement protocols. Another argument could be that such a pooling of data could lead to more robust models, which is also to be examined in the next sections.

### 2.2 Generalizability of Deep Learning Models Across ICU Datasets

Tables 1 and Table 2 present the results for the generalization of the model and training of the target in three deep learning architectures: convolutional neural networks (CNN), InceptionTime and long-short-term memory networks (LSTM). The column labeled “Generalization” denotes model performance when trained on a source domain and directly evaluated on a target domain without additional training. We also report the performance of the model on the target test set when trained with varying proportions (5%, 10%, 20%, 50%, 75%, and 100%) of the target domain training data.

**Table 1.** AUROC performance of models is evaluated from source to target. The deep learning model types include CNN, InceptionTime, and LSTM. Generalization refers to the performance of the source model on the target testing set. The percentages 5%, 10%, 20%, 50%, 75%, and 100% represent the performance achieved when that proportion of the training set is used to train a model (target training) and then evaluated on the target testing set. The pink color indicates that the target training performance is better than the generalization performance. The green color indicates that the target training performance is lesser than the generalization performance.

| Source to Target | Model | Generalization | Target Training |  |  |  |  |  |
| --- | --- | --- | --- | --- | --- | --- | --- | --- |
|  |  |  | 5% | 10% | 20% | 50% | 75% | 100% |
| **MIIV to HiRID** | CNN | 0.676 | 0.567 | 0.679 | 0.675 | 0.699 | 0.750 | 0.748 |
|  | InceptionTime | 0.647 | 0.634 | 0.672 | 0.677 | 0.675 | 0.721 | 0.767 |
|  | LSTM | 0.655 | 0.650 | 0.649 | 0.674 | 0.728 | 0.719 | 0.720 |
| **eICU to HiRID** | CNN | 0.689 | 0.567 | 0.679 | 0.675 | 0.699 | 0.750 | 0.748 |
|  | InceptionTime | 0.661 | 0.634 | 0.672 | 0.677 | 0.675 | 0.721 | 0.767 |
|  | LSTM | 0.647 | 0.650 | 0.649 | 0.674 | 0.728 | 0.719 | 0.720 |
| **HiRID to MIIV** | CNN | 0.665 | 0.523 | 0.686 | 0.718 | 0.751 | 0.736 | 0.750 |
|  | InceptionTime | 0.627 | 0.495 | 0.649 | 0.697 | 0.758 | 0.758 | 0.755 |
|  | LSTM | 0.642 | 0.589 | 0.687 | 0.719 | 0.735 | 0.748 | 0.736 |
| **HiRID to eICU** | CNN | 0.610 | 0.624 | 0.630 | 0.617 | 0.679 | 0.696 | 0.702 |
|  | InceptionTime | 0.597 | 0.612 | 0.601 | 0.648 | 0.691 | 0.693 | 0.703 |
|  | LSTM | 0.614 | 0.634 | 0.615 | 0.640 | 0.679 | 0.691 | 0.701 |

**Table 2.** Normalized AUPRC performance of models is evaluated from source to target. The deep learning model types include CNN, InceptionTime, and LSTM. Generalization refers to the performance of the source model on the target testing set. The percentages 5%, 10%, 20%, 50%, 75%, and 100% represent the performance achieved when that proportion of the training set is used to train a model (target training) and then evaluated on the target testing set. The pink color indicates that the target training performance is better than the generalization performance. The green color indicates that the target training performance is lesser than the generalization performance.

| Source to Target | DL Model | Generalization | Target Training |  |  |  |  |  |
| --- | --- | --- | --- | --- | --- | --- | --- | --- |
|  |  |  | 5% | 10% | 20% | 50% | 75% | 100% |
| **MIIV to HiRID** | CNN | 1.979 | 1.186 | 2.242 | 2.067 | 2.177 | 2.791 | 2.732 |
|  | InceptionTime | 1.622 | 1.715 | 2.202 | 2.104 | 2.143 | 2.450 | 2.987 |
|  | LSTM | 2.096 | 1.861 | 1.749 | 1.978 | 2.456 | 2.673 | 2.286 |
| **eICU to HiRID** | CNN | 1.944 | 1.186 | 2.242 | 2.067 | 2.177 | 2.791 | 2.732 |
|  | InceptionTime | 1.850 | 1.715 | 2.202 | 2.104 | 2.143 | 2.450 | 2.987 |
|  | LSTM | 1.702 | 1.861 | 1.749 | 1.978 | 2.456 | 2.673 | 2.286 |
| **HiRID to MIIV** | CNN | 1.823 | 1.204 | 2.107 | 2.130 | 2.376 | 2.622 | 2.672 |
|  | InceptionTime | 1.573 | 1.041 | 1.770 | 1.875 | 2.714 | 2.424 | 2.540 |
|  | LSTM | 1.664 | 1.289 | 2.270 | 2.000 | 2.270 | 2.473 | 2.479 |
| **HiRID to eICU** | CNN | 1.615 | 1.476 | 1.554 | 1.447 | 1.849 | 2.033 | 2.132 |
|  | InceptionTime | 1.432 | 1.443 | 1.362 | 1.544 | 2.073 | 1.957 | 2.128 |
|  | LSTM | 1.539 | 1.518 | 1.353 | 1.597 | 1.852 | 1.962 | 2.072 |

Overall, these results confirm that pre-trained models offer clear advantages primarily in scenarios with limited target-domain data. Specifically, with only 5% of target data available, using pre-trained models consistently yielded better performance compared to training solely on the limited target dataset. Among the architectures evaluated, CNN models demonstrated slightly superior generalization capabilities, performing robustly across diverse datasets. Both AUROC and normalized area under the precision-recall curve (nAUPRC) steadily improved as more target-domain data became available, eventually reaching a plateau. Although MIIV and HiRID reached comparable performance levels in 100% data use, the eICU dataset consistently exhibited lower performance by at least 0.05 AUROC and nAUPRC, indicating structural or demographic differences inherent to the patient population of the eICU or data collection methods, thus limiting achievable performance.

Dataset-specific observations are provided below: *HiRID as target domain —* Models trained or adapted for HiRID exhibited significant performance variability across different training set sizes. With only 5% of target data available, pre-trained models notably outperformed models trained exclusively on the small subset, with AUROC improvements of up to 0.12 for CNN models (particularly when sourced from the eICU). Notably, models pre-trained on eICU data generalized better than those sourced from MIIV, indicating higher robustness from eICU-trained models. As the target subset expanded to 10% and 20%, the AUROC values for target training and generalization converged, although nAUPRC remained higher for models trained directly on HiRID data. Once 50% or more of HiRID data became available, (*in-domain* target training consistently outperformed generalization. Thus, pre-trained models are beneficial in highly data-limited scenarios (up to approximately 20% of the target data), but direct target training becomes more advantageous once moderate amounts of HiRID data (over 50%) are available. *MIIV as target domain —* All three architectures showed consistent AUROC trends, although nAUPRC exhibited some variability. With limited target data (5%), pre-trained models outperformed direct training on MIIV alone. However, as target data availability increased beyond 10%, direct training in the larger MIIV subset exceeded the generalization of source-trained models. In particular, AUROC performance plateaued around the 50% data threshold, while nAUPRC continued to improve slightly with additional data. This discrepancy suggests that the basic discriminative capability (AUROC) stabilizes earlier, whereas precision recall metrics continue to improve incrementally with additional data, underscoring the importance of data volume for precision recall outcomes. *eICU as target domain —* On the contrary, eICU exhibited relatively stable performance curves with minimal variability across different subset sizes. At the smallest training fraction (5%), direct training provided marginal AUROC improvements over generalization from pre-trained models. As training subsets expanded, performance plateaued markedly around two levels: an initial plateau at 5 to 20% and a second at 50 to 100% of available data. The narrower gap between these plateaus in the eICU compared to HiRID and MIIV suggests greater inherent data set diversity (reflecting its multicenter nature) or structural characteristics that make it easier to robustly model the prediction of sepsis, even with limited samples.

Collectively, these results confirm that models pre-trained on source domains yield substantial benefits in highly data-constrained target scenarios but become progressively less advantageous as more target-domain data are available. Furthermore, observed differences due to clinical practices, demographics of patients, and documentation protocols, especially in the eICU, underscore the importance of accounting for dataset-specific factors when leveraging external datasets for prediction of sepsis in ICU settings.

### 2.3 Systematic Comparison of the Five Deployment Strategies

In Section 2.2, we discussed how generalization and target training compare to each other. Next, we systematically evaluated the other four approaches to training or adapting deep learning models: fine-tuning, retraining, supervised domain adaptation (DA), and fusion training, across four source-to-target dataset combinations (Table 5). We also compare these performance against generalization (baseline) and target training performance, discussed earlier. The goal is to identify how effectively each approach leverages target data under varying sizes and conditions, since we already established that these datasets have considerable distribution shifts. Figure 9 shows the AUROC and nAUPRC box plots, illustrating the performance of each method by the combination of source-to-target and the size of the target data set (small, medium, large, as described in Figure 7). In all these figures, darker tone colors 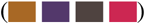 represent DA methods, mid tone colors 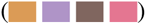 indicate standard transfer learning approaches, and lighter tone colors 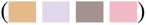 indicate fusion training. Moreover, each source-to-target combination is assigned a distinct color. For example, MIIV to HiRID in 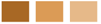, HiRID to MIIV in 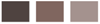, eICU to HiRID in 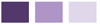, and HiRID to eICU in 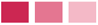.

**Figure 7.**
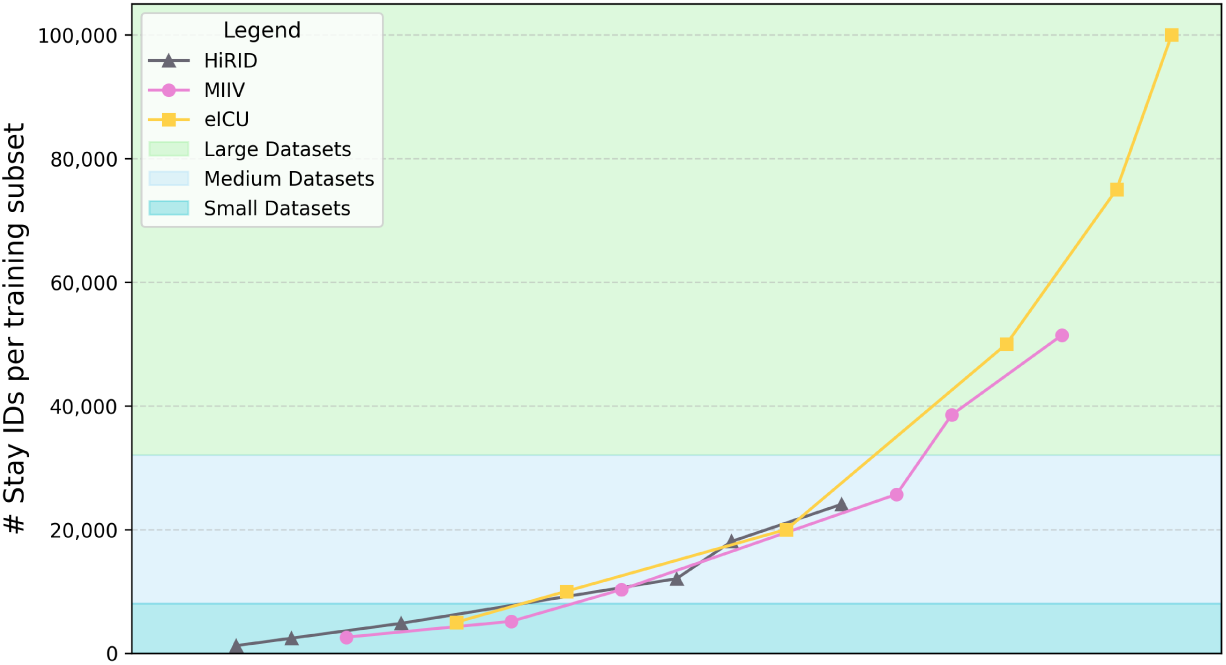
Visualization of dataset size groupings across HiRID, MIIV, and eICU training subsets. The figure illustrates the 18 training subsets (six subsets for each dataset: HiRID, MIIV, and eICU) categorized into small, medium, and large size groups based on the number of stay_ids. The subsets are arranged in ascending order of size, with each point representing a specific subset percentage (5%, 10%, 20%, 50%, 75%, and 100%) within its respective dataset.

**Figure 8.**
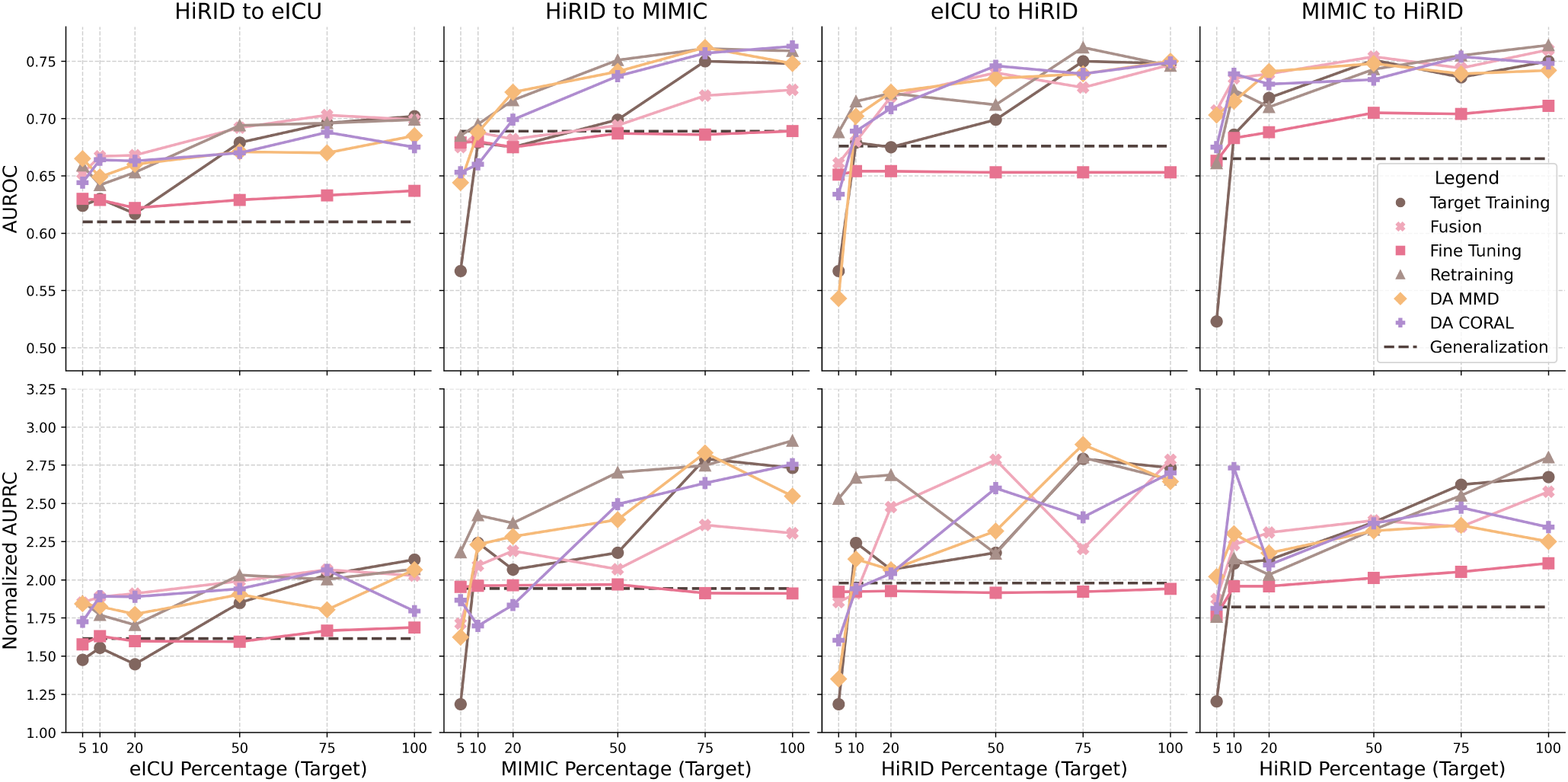
Comparison of CNN model performance across different stratefies, for different source to target pairs. Each subplot shows the AUROC (top row) and normalized AUPRC (bottom row) as a function of the target dataset percentage used in the training process. The dashed brown line represents the Generalization baseline (performance of the source model in the target domain). Colored solid lines correspond to different training strategies, including Target Training, Fusion, Fine Tuning, Retraining, DA MMD, and DA CORAL.

**Figure 9.**
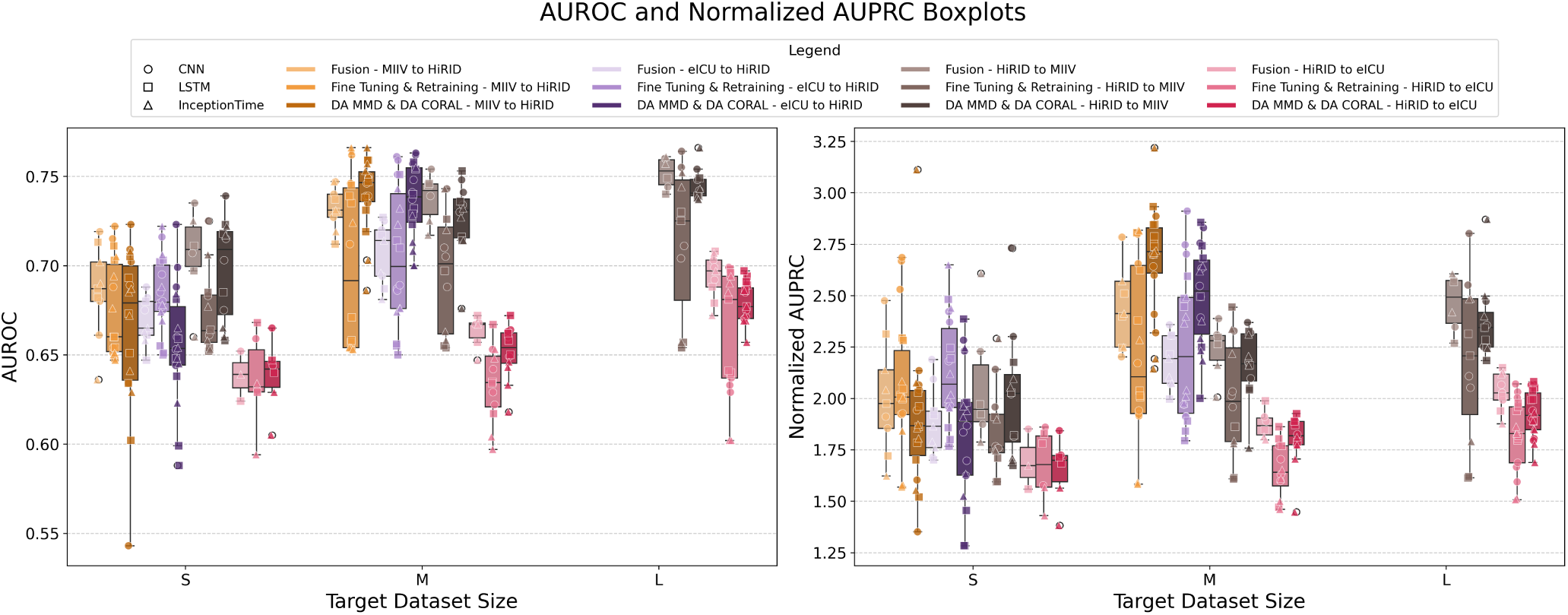
Joint DL models’ performances grouped by source-to-target combination, target dataset size (small, medium and large) and transfer learning techniques.

First, if we consider the scenario where only a pre-trained model from the source domain is available, along with incrementally increasing amounts of target domain data (small [S], medium [M], and large [L]), the feasible strategies include generalization, fine-tuning, retraining, and direct target training. The results comparing these approaches across four distinct combinations of source-to-target datasets, using a CNN model, are shown in Figure 8 (Figures 14-17 provide a detailed breakdown of the results, even for other model types). As established in Section 2.2, except for scenarios involving minimal target training data (approximately 5%-10%), direct target training generally outperforms the straightforward generalization approach. This trend is clearly evident in the results presented. Comparing standard transfer learning methods (fine-tuning or retraining) with direct target training reveals that CNN-based models that utilize retraining consistently outperform direct target training in the overwhelming majority of cases. Similar patterns are observed for InceptionTime and LSTM architectures, although to a lesser extent. This finding suggests that, irrespective of the quantity of available target domain data, retraining a pre-trained source model typically yields superior model performance compared to training from scratch on the target domain alone.

Next, if we consider the scenario in which both the data and the model from the source domain are available alongside incremental target domain data, all six strategies become viable. In a more constrained scenario, where only the source data (but not a pre-trained model) are accessible along with the target data, the options reduce to direct target training, fusion training, and supervised domain adaptation. The results of these setups are presented in Figure 9 as boxplots and are detailed in Figure 8. The boxplot results for both AUROC and nAUPRC indicate that, for medium-scale target datasets, supervised domain adaptation techniques clearly outperform more simplistic methods, such as fusion training or standard transfer learning. Even for smaller target datasets, supervised domain adaptation yields the highest peak performance across all models, particularly evident for CNN architectures, as illustrated in Figure 13 in the Appendix. Moreover, regardless of the size of the source dataset, domain adaptation consistently exhibits lower performance variability (illustrated by shorter boxes in the plots), indicating a more stable and robust generalization. In small data contexts, supervised domain adaptation outperforms standard transfer learning predominantly when the source domain dataset is sufficiently large (such as MIIV or eICU). In contrast, standard transfer learning approaches demonstrate significantly greater variability (longer boxes), underscoring their sensitivity to differences in dataset size and composition.

Further analysis based on nAUPRC emphasizes a clear advantage when leveraging larger or medium-sized source datasets, such as eICU (represented in purple) or MIIV (in orange). Models trained on these datasets generally outperform those initialized from smaller source datasets (shown in gray and pink). Furthermore, the characteristics of specific data sets have a significant impact on model performance. For example, models trained on the HiRID source data set achieve better results when applied to the MIIV target data set compared to the eICU target data set, regardless of target subset size. This observation implies that the compatibility and similarity of the data set, beyond the mere quantity of data, significantly affect the performance. Furthermore, comparisons among the three deep learning (DL) architectures highlight the interaction between model design and data set size. As illustrated in Figure 13 of the Appendix, the CNN and LSTM architectures generally follow the observed trends, with CNN showing slightly lower variance and consistently outperforming LSTM, demonstrating robustness across various domain changes. InceptionTime exhibits the greatest performance variability but performs notably well for small target datasets, particularly when paired with larger source datasets (MIIV or eICU).

To summarize all these results across different sources-target domain pairings, we created a ranking as given in Figure 10, where for each target domain dataset size (small, medium and large), the performance of the approaches is averaged and ranked from highest to lowest, while also showing differences between different approaches using colored brackets. The results here confirm and synthesize our findings that for all models (Figure 10A, Figure 10C) and even for CNNs specifically (Figure 10B and Figure 10D), re-training is the superior method when the target dataset size is small, followed by fusion training. For medium-sized target domains, DA MMD and DA CORAL were superior. For large target domains, CNN models worked the best with retraining, whereas the highest performance when all models were combined was achieved with Fusion training (in terms of AUROC) and target training (in terms of nAUPRC). Furthermore, as the brackets indicating statistical significance show, highest performing techniques (retraining, DA, fusion) showed statistically significant performance improvements over commonly explored approaches such as fine-tuning and target training, particularly for small and medium target domains.

**Figure 10.**
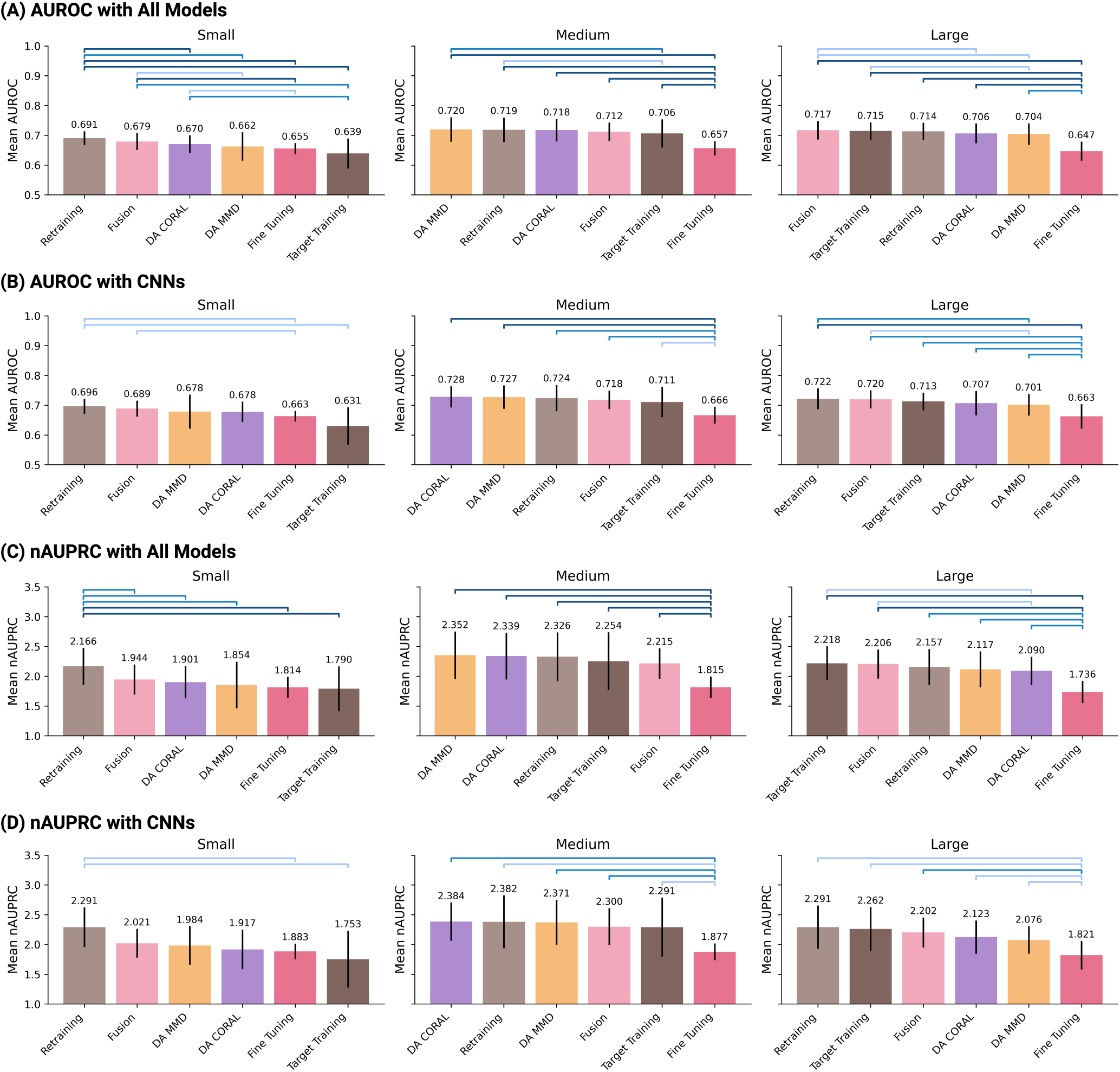
Rankings of strategies for models based on AUROC and nAUPRC. Colored brackets indicate statistically significant differences between pairs of methods: adjusted p-value *<* 0.05: 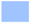; adjusted p-value < 0.01: 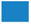; adjusted p-value < 0.001: 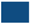. **(A)** AUROC for all models combined; **(B)** AUROC for CNNs only, the generally top-performing model; **(C)** nAUPRC for all models combined; **(D)** nAUPRC for CNNs only, the generally top-performing model.

**Figure 11.**
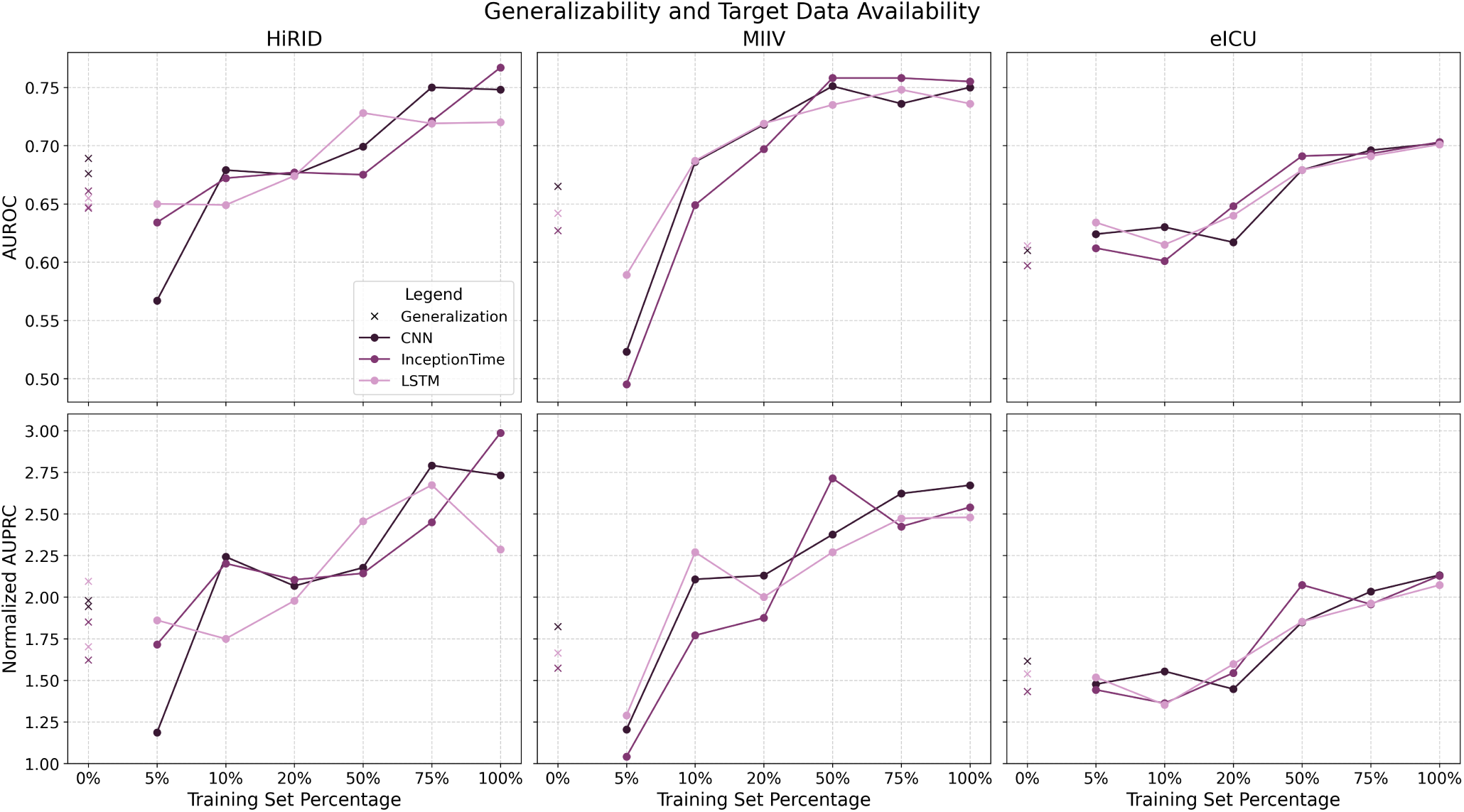
Generalizability Across ICU Datasets and Relation Between Target Data Availability and AUROC and normalized AUPRC Performances. Three columns represent results for the three target domains HiRID, MIIV, and eICU. Generalization refers to how well models trained in source domains, performed for the target domain. The results for all CNN, InceptionTime, and LSTM are shown.

In summary, these findings suggest that supervised domain adaptation (e.g. MMD or CORAL) typically represents a superior strategy for medium-target datasets compared to fusion training, generalization, or direct target training. The size and compatibility of the source dataset critically influence the outcomes, with larger sources (e.g. eICU to HiRID) providing better overall performance. In general, our analysis underscores the importance of tailoring model training and deployment strategies, including the choice of transfer learning methods and DL architectures, to specific characteristics of the source and target data sets, thus maximizing the accuracy and utility of sepsis prediction in deployment. Relying on fine-tuning might not always be the best approach, although it has been the go-to approach examined in previous literature.

## 3 Discussion

Our findings have important implications for the deployment of AI sepsis prediction models in real-world ICU settings. First, we have shown that distribution shifts between hospitals are not just a theoretical concern, but a practical reality that can significantly impact model performance. For clinicians, data scientists, and hospital IT leaders, this means that an accurate sepsis early warning model developed at a larger center might not work as well on their patient population without adjustments. This study reinforces the need for local validation of any model before clinical use; if a model is trained elsewhere, it should be tested on local data and expect some drop in metrics due to the changes we quantified. The good news is that we also demonstrate solutions to overcome these shifts.

Domain adaptation provides a tangible approach to improve the generalizability of the model across different ICUs, even with a small amount of labeled target data. In practical terms, a hospital adopting an externally developed sepsis model can apply domain adaptation by using a modest set of its own data (perhaps a few months of retrospective data) to fine-tune the model in a domain-aware way. For example, suppose that Hospital B wants to use a hospital-acute sepsis model. Rather than training a new model from scratch (which could take years of data collection), Hospital B could use Hospital A’s data as well and train on a sample of 1000 cases of sepsis and non-sepsis from Hospital B, while using a DA technique to ensure that the model’s internal representations adjust to fit Hospital B’s data patterns. Our results suggest that this process can produce a model nearly as good as if Hospital B had years of its own data to train with. This has huge time and cost benefits; it accelerates the availability of AI tools in settings that lack big data resources. Furthermore, because the domain adaptation uses source data during training, the adapted model in Hospital B still benefits from the larger dataset of Hospital A (for general sepsis aspects), but also fits the specific profile of Hospital B (for differences, such as laboratory value distributions or patient demographics).

From a clinical perspective, the ultimate measure of success is whether these models actually improve patient outcomes by requiring earlier or more accurate interventions.A model that performs poorly in a new environment risks generating false alarms or missed detections, undermining clinician trust and potentially compromising patient safety. By mitigating performance degradation, domain adaptation can ensure that predictive accuracy remains high in the new setting, preserving the positive impact on patient care. For example, if without adaptation the model sensitivity to sepsis in Hospital B had dropped, clinicians might miss the early window to treat sepsis in some patients; with adaptation that restores sensitivity, these cases can be caught. Likewise, specificity might be maintained, avoiding alarm fatigue from too many false alerts in the new unit. Basically, DA contributes to safer and more reliable AI. We also found that certain model architectures (such as CNNs) are inherently more stable across domains, suggesting that choosing the right model design is part of making an AI tool robust for deployment. Simpler models that generalize well may be preferable to complex ones that achieve slightly higher accuracy in one setting but fail to transfer. Clinicians and engineers who collaborate on AI should consider this trade-off: a model that is 2% less accurate in one hospital but portable in many hospitals may have a greater overall impact on patient outcomes than a model that is highly optimized in one place only.

Another practical insight is our guideline on when to update a model with local data and when it might be unnecessary. For example, if a hospital is implementing an ICU sepsis model and has virtually no local training data, our results suggest using the model as-is might yield suboptimal performance, but collecting even a small amount of data (a few hundred cases) to adapt the model will produce large gains. On the other hand, if a hospital has a very large trove of its own ICU data (tens of thousands of cases with results), they might consider training their own model or doing extensive retraining, as the marginal benefit from external data is small. Many real-world scenarios will fall between moderate data availability. For those, our work indicates that joint training with domain adaptation is likely the best route, rather than pure fine-tuning. Hospital systems that have multiple sites could use a domain adaptation strategy to create a network of models: a central model plus adapted versions for each site, sharing knowledge but tuned to local peculiarities.

We also highlight that simply merging data from different sites into one large training set is not necessarily a panacea. Although having more data is generally beneficial, our analysis of distribution shifts shows that without explicitly handling those shifts, a merged model may still underperform in each constituent domain. For example, it could have a calibration problem: perhaps systematically overpredict risk in one hospital and underpredict in another if data from one site dominates training. Clinically, this could mean that one hospital’s clinicians receive too many alerts and another too few. A smarter approach might be to merge the data but include domain adaptation or stratification in the modeling process (e.g., domain-specific layers or parameters for each hospital). Our results on the benefits of DA hint that such an approach would outperform a naive pooled model. This is an area for future exploration (and in fact, we suggested ‘fusion training’ with careful consideration as future work).

Lastly, from a clinical governance standpoint, our study underscores the importance of data standards and sharing. The fact that we had to harmonize three datasets and still found so many differences highlights how variable healthcare data can be. If in the future more hospitals adopt common data standards (for example, using common unit measurement and documentation practices), distribution changes might be lessened, making generalization easier. However, given the reality of the differences between patients and practice, some level of change will always persist. Therefore, having methods like domain adaptation in the toolbox can facilitate the translation of medical AI innovations from one setting to another. This is analogous to how a drug developed in one population might need dose adjustments in another – an AI model might need a ‘dose adjustment’ in the form of retraining or adaptation when moving to a new ICU population.

### Limitations

Although this study provides important information, it also has several limitations that point to areas for further caution and research. First, our work is retrospective in nature. We evaluated model performance using historical ICU data from three databases and, while we simulated the effect of deploying a model in a new domain, we did not prospectively implement these models in real ICU workflows. Thus, factors such as availability of real-time data, integration with clinical systems, and human-in-the-loop considerations were not tested. Some practical issues (e.g. missing data that arrive late or clinicians reacting to model outputs) could affect the performance of a model deployed in ways not captured here. Future work should include prospective or simulation studies in which an adapted model is put into practice to confirm the benefits on live data and patient management.

Second, we examine only a subset of possible domain-adaptive techniques. We chose MMD and CORAL for their simplicity and proven performance in other domains, but there are more advanced methods. For example, adversarial domain adaptation (such as training a model to fool a domain discriminator, as done in Domain-Adversarial Neural Networks)^16^ could potentially learn even more invariant features. Generative approaches or normalizing flows could explicitly transform data from one domain to another distribution. Furthermore, recent techniques in transfer learning, such as fine-tuning with discriminative learning rates, or parameter-efficient transfer (like freezing most of the model and only learning small adapter modules), were not explored and could be beneficial in this context. Our results open the door to testing these methods: given that we have established sepsis prediction as a problem domain where adaptation helps.

Third, our analysis of distribution shifts was somewhat high-level. We identified and quantified differences in feature distributions, but we did not delve deeply into the root causes of each difference, and also the clinical implications of these findings. For example, we note that the neutrophil values of the band ranged from 0 to 100% in one dataset versus 0 to 10% in others; this could be due to different laboratory reports (perhaps one reports band forms as a percentage of WBC, another as an absolute count or different lab standards). We did not attempt to standardize such features beyond what we did initially; perhaps some differences could be eliminated by better data pre-processing (unit conversion, outlier handling) if one knows the cause. We also did not explore concept drift or label differences, and we assumed that the Sepsis-3 criteria were applied uniformly, but it is possible that the way sepsis onset is identified in each data set had subtle differences (e.g., exactly how infection suspicion was determined from the chart data). If the sepsis labels themselves are slightly inconsistent, that could introduce another kind of shift (target shift or label shift), which we did not explicitly address. Our focus was on covariate shift (features), and we treated the prediction task as identical across domains. In reality, one limitation is that sepsis detection criteria might vary or have noise; something that future work could investigate by, say, using an alternate sepsis definition or physician adjudication to see if results hold.

Finally, this study was limited to three particular datasets (MIMIC-IV, eICU, HiRID). All are large, high-quality clinical research databases from tertiary ICUs. They may not capture the full spectrum of ICU types (for example, community hospital ICUs, pediatric ICUs, etc.) were not in scope. Thus, the shifts we observed (geography and certain practice differences) might be different in other settings (for example, a shift due to temporal changes or due to a different EHR system). The general principle should still apply, but the specific differences in features and the magnitude of the issues might vary. We encourage validation of our findings on other datasets as they become available (e.g., there are emerging multicenter ICU datasets from other countries). Furthermore, we only looked at adult sepsis; pediatric sepsis prediction is another area where data are more scarce and changes (between children’s hospitals) could be significant, where adaptation could be even more crucial there.

## 4 Methods

### 4.1 Dataset Harmonization and Pre-Processing

We used three open, high-resolution ICU databases to ensure a diverse representation of patient populations and clinical practices. MIMIC-IV (a single-center US ICU database)^17^, eICU Collaborative Research Database (a multicenter US ICU database)^18^, and HiRID (a single-center Swiss ICU database)^19^. Collectively, these databases encompass more than 216,000 stays in the ICU, including more than 10,800 patients who developed sepsis during their ICU course. All three data sets were filtered to include adult stays in the ICU and annotated with sepsis outcomes based on Sepsis-3 criteria (which require suspicion of infection plus organ dysfunction). We harmonized variable definitions and units across databases using a rigorous pre-processing pipeline. This involved mapping the raw features of each dataset to a common set of clinical variables (e.g., vital signs such as heart rate, lab measurements such as creatinine) with consistent units and naming. We focus on 48 time-varying physiological features available in all three sources, along with relevant demographics. Continuous signals were resampled or summarized into hourly intervals to allow alignment. Outliers were handled by capping physiologically implausible values, and each time series was filled forward and imputed as needed to handle missingness. To ensure label consistency, we applied the same sepsis onset detection logic (based on the increase in the Sequential Organ Failure Assessment score and the clinical documentation of infection) uniformly to all datasets, verifying that the incidence rates matched published values for each database.

For data set harmonization, we closely followed the methodology outlined in YAIB^15^. By adopting this framework, we ensured consistency and reproducibility in the dataset haromization process, without having to make individual decisions regarding data preprocessing, cohort definitions, or task evaluation. Hence, the HiRID, MIIV and eICU datasets were harmonized, ensuring that all datasets shared the same set of characteristics and structure. Specifically, the harmonized data sets included four static features and 48 dynamic features (7 vitals, 39 lab tests, urine output, and fraction of inspired oxygen). These features were available an hourly basis from day 1 to day 7 of the ICU stay. To define sepsis, I adhered strictly to the Sepsis-3 criteria, including the length of antibiotic treatment and body fluid culture to characterize suspicion of infection. In addition, to confirm sepsis, antibiotics were required to be administered continuously for at least three days. For labeling purposes, the sepsis label was set to “True”, six hours before the confirmed onset of sepsis. This allowed the model to account for early warning signs of sepsis, hence making the task we attempt a sepsis prediction task, six hours in advance. Regarding the temporal aspect, a maximum of 13 hours with the “True” label was kept after the onset of sepsis, to make sure that too distant data points from the onset are considered, making the inference easier and biased. The total number of stay_ids present in each of the datasets after harmonization as well as the total number of stay_ids which developed sepsis are presented in Table 3.

**Table 3.** Number of Stay_ids within the Harmonized Datasets.

| Dataset | eICU | MIMIC-IV | HiRID |
| --- | --- | --- | --- |
| Total number of stay_ids | 123,413 | 63,425 | 29,698 |
| Total number of stay_ids with sepsis | 5,638 | 3,321 | 1,858 |
| % of stay_ids with sepsis | 4.568% | 5.236% | 6.256% |

To further preprocess the harmonized ICU datasets, several steps were taken to ensure that the data were clean, standardized, and suitable for training DL models. Each data set was divided into three subsets according to the timestamp: 80% for training, 10% for testing, and 10% for validation. This division ensured that the model had ample data for training while also having separate data to evaluate its performance during training (validation) and after training (testing). Importantly, all data were stratified by stay_id and label, preventing any data leakage and ensuring that each split contained a balanced representation of patient stays across all three subsets. The number of stay_ids present in each of the three splits is displayed in Table 4.

**Table 4.**
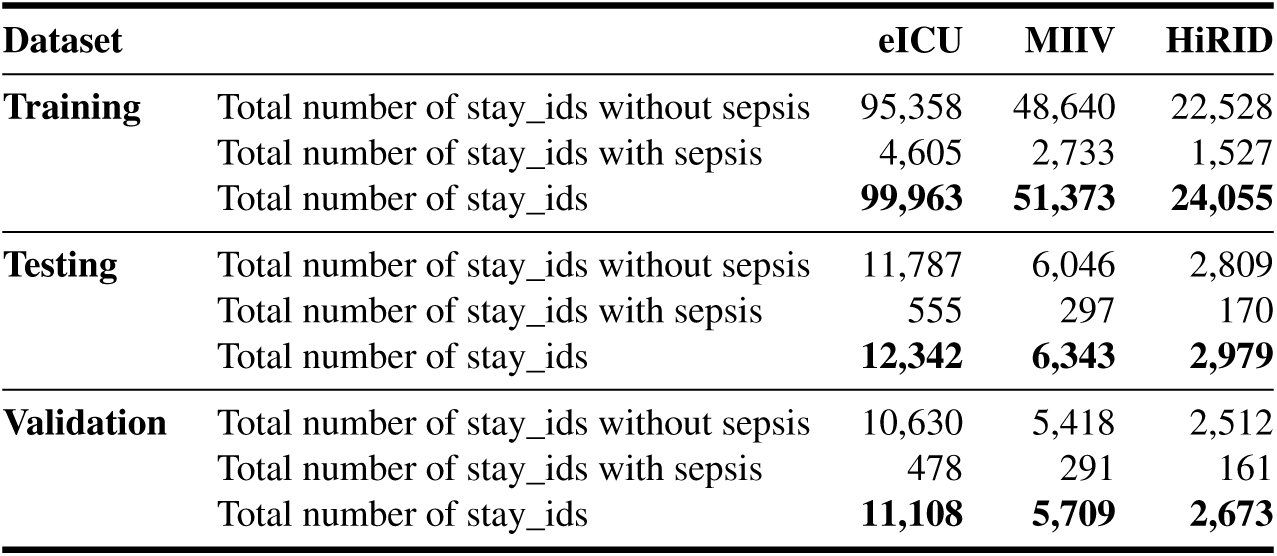
Training, Testing and Validation Splits.

| Dataset |  | eICU | MIIV | HiRID |
| --- | --- | --- | --- | --- |
| **Training** | Total number of stay_ids without sepsis | 95,358 | 48,640 | 22,528 |
|  | Total number of stay_ids with sepsis | 4,605 | 2,733 | 1,527 |
|  | Total number of stay_ids | **99,963** | **51,373** | **24,055** |
| **Testing** | Total number of stay_ids without sepsis | 11,787 | 6,046 | 2,809 |
|  | Total number of stay_ids with sepsis | 555 | 297 | 170 |
|  | Total number of stay_ids | **12,342** | **6,343** | **2,979** |
| **Validation** | Total number of stay_ids without sepsis | 10,630 | 5,418 | 2,512 |
|  | Total number of stay_ids with sepsis | 478 | 291 | 161 |
|  | Total number of stay_ids | **11,108** | **5,709** | **2,673** |

Given the high frequency of missing values, particularly in dynamic features (i.e., lab results), some strategies were used to handle missing values. Initially, missing values in dynamic features were flagged, creating an additional binary column for each feature that indicated whether a value was missing. This resulted in every dynamic feature having two corresponding columns: the original value and the missingness flag. After flagging, the missing values were re-filled, which means that they were replaced with the last available measurement for the same patient during their ICU stay. If no prior measurement was available for a feature, the missing value was filled with the mean of that feature, calculated solely from the training set to avoid data leakage. This mean imputation was determined before any forward filling was applied, ensuring a consistent and unbiased approach throughout the data set. To prepare the dynamic features for model training, they were standardized using the StandardScaler function from the Scikit-learn library. This process scaled each feature to have zero mean and unit variance. By standardizing the features, all variables were brought onto a comparable scale, ensuring that no single feature disproportionately influenced model training. Importantly, the scaling parameters (mean and variance) were computed using only the training set and then applied to both the validation and testing sets.

For model experimentation, the training set was further split into six progressively larger subsets: 5%, 10%, 20%, 50%, 75%, and 100%. In constructing these subsets, additional stay_ids were added sequentially to each larger subset. For example, the 10% subset included all stay_ids from the 5% subset plus 5% new stay_ids. Similarly, the 20% subset contained all stay_ids from the 10% subset, with an additional 10% of new stay_ids, and so on. This incremental addition ensured that each larger subset was an expansion of the previous one, preserving continuity in the dataset composition while enabling an evaluation of the impact of data size on performance. Importantly, each subset was stratified by stay_id and label to maintain a balanced representation of ICU stays across all training subsets. Evaluating different dataset sizes is particularly important given that this is the sequential nature in which data are collected and stored in ICUs, in the real world. Moreover, to capture the temporal dynamics of patient data, the data was segmented into 6-hour windows based on the stay_id of each patient. After extraction, the time and stay_id columns were removed from the datasets.

### 4.2 Training Deep Learning Models

To evaluate the performance of DL models on sepsis ICU datasets, we implemented three model architectures (see Appendix A): Convolutional Neural Networks (CNN), InceptionTime, and Long-Short-Term Memory (LSTM) networks. These model architectures were chosen because they have shown decent performance for time series modeling tasks, including predictive tasks in the ICU. Model performance was assessed using two primary metrics: the Area Under the Receiver Operating Characteristic Curve (AUROC) and the normalized Area Under the Precision-Recall Curve (nAUPRC). The nAUPRC was selected for its ability to address significant class imbalance, as it adjusts the raw AUPRC by dividing it by the baseline, defined as the proportion of positive class samples (“True” labels) in the training dataset:

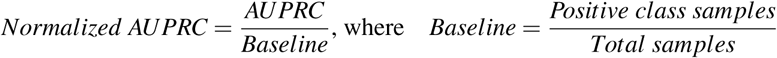

This normalization accounted for the inherent difficulty posed by class imbalance. An nAUPRC of one reflected baseline performance (random guessing), while values greater than one indicated better than baseline performance.

Models were trained using a batch size of 32, binary cross-entropy with logit loss (BCEWithLogitsLoss) as the loss function, with positive samples weighted to address class imbalance. Training employed early stopping, terminating if the validation loss kept increasing for five consecutive epochs. The best performing models, determined by the lowest validation loss, were saved. All architectures used the Leaky ReLU activation function to mitigate vanishing gradient problems for negative activations. Additionally, the same random seed was set across all models to ensure reproducibility. For the CNN model, we used the stochastic gradient descent (SGD) optimizer with a learning rate of 0.01, a momentum factor of 0.85, and a weight decay of 0.0001. A learning rate scheduler reduced the learning rate by a factor of 0.9 if the validation loss plateaued for five consecutive epochs, with a minimum learning rate of 0.001. Training was carried out for up to 150 epochs. The LSTM model was optimized using Weighted Adam (AdamW), with an initial learning rate of 0.001. A learning rate scheduler decreased the learning rate by a factor of 0.85 if the validation loss failed to improve for five consecutive epochs, with a minimum learning rate of 0.0001. The training process spanned 120 epochs. Lastly, for the InceptionTime model, a depth of 12 modules was used to extract hierarchical temporal features^20^. For this, we used the InceptionTime model architecture by Fawaz et al.^21^. The optimizer was AdamW with a learning rate of 0.01. A learning rate scheduler adjusted the learning rate by a factor of 0.9 based on stagnation of the validation loss, with a minimum value of 0.001. Training was limited to 60 epochs due to the model’s higher computational costs. Again, with early stopping, none of the models reached the maximum number of epochs we set, when starting the model training.

### 4.3 Experimental Setup

#### 4.3.1 Existence and Significance of Distribution Shifts

To explore the presence and significance of covariate distribution shifts between ICU data sets, we performed a comparative analysis of the three data sets. Since the MIIV and eICU datasets originate in the United States, while the HiRID dataset comes from Switzerland, we also examined whether these shifts were more pronounced when comparing the datasets from the two countries. The analysis focused on the three dataset pairings: HiRID-MIIV, HiRID-eICU, and MIIV-eICU. The data sets analyzed were the unprocessed harmonized versions, which means that they retained missing values and had not undergone preprocessing steps such as forward filling or imputation (see Section 4.1). This approach allowed for a more accurate assessment of the raw distributional differences between datasets.

To quantify distribution shifts, we performed a statistical analysis. For each dynamic feature and stay_id, we calculated descriptive statistics including the mean, variance, minimum, and maximum values, resulting in a total of 192 features (48 *dynamic f eatures ×* 4 *descriptive statistics*). To assess the statistical significance of the shifts, we performed pairwise comparisons of the three dataset pairings using the two-sample Kolmogorov-Smirnov (K-S) test, with a Benjamini/Hochberg correction to account for multiple testing. Features with p-values less than 0.01 were considered significant. In addition to the statistical analysis, the importance of the distribution shifts were quantified by measuring effect sizes using the absolute Cohen’s-d. The shifts were classified as small effect size (0.2-0.5), medium effect size (0.5-0.8), or large effect size (greater than 0.8). This dual approach, including statistical testing and effect size estimation, aimed to provide a comprehensive view of the magnitude and relevance of the observed shifts, particularly in the context of datasets from different countries.

To visually explore feature distributions, we created violin plots of the dynamic features of each data set. For added granularity, we stratified each dataset based on patient stay_ids into two subgroups: (1) stays where patients developed sepsis, with distributions plotted separately for the periods before and during sepsis, and (2) stays where patients did not develop sepsis, focusing solely on the negative label distribution.

#### 4.3.2 Generalizability of Models Across ICUs

The dataset combinations presented in Table 5 were used consistently in the next set of experiments. These combinations were designed to evaluate the generalizability of sepsis detection models across ICU datasets and assess the impact of training data availability on model performance.

**Table 5.**
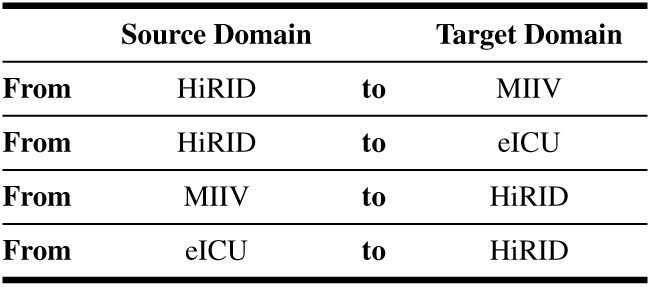
Source and Target Datasets Combinations.

The first set of experiments examined whether models trained on a source domain could maintain robust performance when tested on an unseen target domain without further training. The three DL architectures—LSTM, CNN, and InceptionTime— were trained using 100% of the source training dataset and evaluated on the target testing dataset. This setup allowed for a direct assessment of model generalizability under distribution shifts.

In the second step, the focus shifted to the influence of target domain training data on model performance. Models were trained exclusively using progressively larger subsets of the target training data, comprising 5%, 10%, 20%, 50%, 75%, and 100% of the target dataset. Performance was evaluated on both the target testing and validation sets. This analysis compared the effectiveness of pre-trained models (without additional training) against newly trained models across varying data availability. The goal was to determine whether a pre-trained model could outperform newly trained models, even when only limited target data were available.

#### 4.3.3 Experiments on Different Deployment Strategies

This section evaluates techniques to enhance model generalization in the target domain, comparing standard transfer learning approaches (fine-tuning and retraining) with supervised DA methods (MMD and CORAL) and fusion training. Each approach was assessed using target training datasets of varying sizes (5%, 10%, 20%, 50%, 75%, and 100%), while the source training dataset was maintained at 100%. The same four source-to-target dataset combinations (Table 5) were used as in RQ2. The selection of the model was based on the lowest validation loss, with evaluations performed consistently on the target testing data set.

First, standard transfer learning techniques - fine-tuning and retraining - were applied to adapt pre-trained models to the target domain. Fine-tuning involved updating only the final fully connected layers of the pre-trained models, keeping the earlier layers frozen to preserve general feature representations learned from the source domain. This approach helps maintain the model’s ability to extract generalizable features while adapting to the target domain and reduces the risk of overfitting to smaller target datasets^8^. Adjustments were made to the learning rate scheduler to accelerate convergence during training, with factors set to 0.75 for CNN and InceptionTime and 0.7 for LSTM. Apart those, no other changes were made.

In contrast, full retraining updated all weights of the pre-trained models, using them as initialization for training on the target domain. This approach allows the models to adapt more comprehensively to the target dataset while retaining useful representations from the source domain, based on the initialization. To prevent overfitting and ensure stable optimization, optimizer learning rates were reduced, and the learning rate scheduler’s factor was divided by 10 to maintain an optimal ratio with the optimizer. In addition, dropout rates were increased and dropout layers were added to enhance regularization, to avoid overfitting. Specific modifications for each architecture included—CNN: The optimization learning rate was set at 0.001, the probability of dropout increased from 0.5 to 0.6, and an additional dropout layer was added; LSTM: The optimization learning rate was set at 0.0001, and the dropout rates were uniformly increased to 0.6; InceptionTime: The optimization learning rate was set to 0.0001, the probability of dropout increased from 0.3 to 0.7, and a dropout layer was added after each InceptionTime module.

DA, another form the advanced transfer learning strategy, incorporated labeled source and target data to compute a domain alignment loss, which aims to align feature distributions between the two domains. To address differences in dataset sizes, the smaller dataset (source or target) was repeated to match the size of the larger dataset within each epoch. The features were extracted for alignment after the model embedding layers, before the task classifier^22^. The total loss function combined classification losses from the source and target domains with the domain alignment loss scaled by the adaptation weight *λ*, calculated as: *Total Loss* = *Source Loss* + *Target Loss* + (*λ ×DA Loss*). MMD used a Gaussian RBF kernel to align the statistical means of the source and target feature distributions in a shared feature space. We based our code on the implmenet on Lee et al.^23^. Key hyperparameters included a fixed bandwidth length (*f ix*_*sigma* = 1.0), the Gaussian RBF Kernel’s bandwidth parameter (*kernel*_*mul* = 2.0), and the number of Gaussian kernels with different bandwidths (*kernel*_*num* = 5). For CNN, a *kernel*_*mul* = 5.0 was chosen because *λ* -tuning did not yield satisfactory results. The adaptation weights *λ* were tuned in different combinations of the source and target data sets, and the value that provided the best overall performance was selected. *λ* was set to 1 for CNN and LSTM, and 10 for InceptionTime. Moreover, CORAL achieved alignment by minimizing the distance between the covariance matrices of the source and target feature embeddings. The implementation of CORAL was adapted from Github of the Institute of Computational Perception^22^. Adaptation weights *λ* were adjusted for each architecture and set to 1,000 for CNN and 10,000 for LSTM and InceptionTime. No additional hyper-parameters were tuned.

With these experiments, we also wanted to understand the effect of source and target domain’s training dataset size, on the model performance. In the previous experiments, models were trained on datasets of varying sizes. When using 100% of the training data, the total number of stay_ids was approximately 24K for HiRID, 51K for MIIV, and 100K for eICU (see Figure 7). To systematically analyze whether the size of both source and target datasets influences a model’s generalization ability, as well as whether specific transfer learning methods perform better under certain conditions, we categorized the datasets into small, medium, and large groups. The training sets for the source domain were classified as follows: HiRID as small, MIIV as medium, and eICU as large datasets. For the target domain training subsets, categorization was based on the number of stay_ids, with small datasets containing up to 8,000 stay_ids, medium datasets containing between 8,000 and 32,000 stay_ids, and large datasets containing more than 32,000 stay_ids. Table 6 maps each training subset to its respective group, while Figure 7 visually illustrates these groupings. In the figure, each dataset (HiRID, MIIV, and eICU) is represented by six points corresponding to the six training subsets (5%, 10%, 20%, 50%, 75%, and 100%). These 18 training subsets (3 datasets *×* 6 subsets) are arranged in ascending order of dataset size for clarity. This categorization allows for a systematic exploration of how dataset size impacts model performance and enables comparison across different training scenarios.

**Table 6.** Training Subsets in each Size Group.

| Dataset | eICU | MIIV | HiRID |
| --- | --- | --- | --- |
| **Small:**<br>0-8,000 stay_ids | 5% | 5%, 10% | 5%, 10%, 20% |
| **Medium:**<br>8,000-32,000 stay_ids | 10%, 20% | 20%, 50% | 50%, 75%, 100% |
| **Large:**<br>≥ 32,000 stay_ids | 50%, 75%, 100% | 75%, 100% | - |

For each target size bin (small, medium, large), we collected every AUROC and nAUPRC value produced in Section 4.3.3. This produced, per deployment strategy, a matched set of 4 source*→*target pairs *×* 3 architectures = 12 scores (for the panels ‘all models’) or 4 *×* 1 = 4 scores when the analysis was restricted to CNNs. Within a bin we averaged those scores and assigned ordinal ranks (1=best, 6=worst; ties were broken by retaining additional significant digits). To test whether the leading strategy was meaningfully better than its competitors, we applied a paired Wilcoxon signed rank test to the matched score vectors of every pair of strategies, correcting *p* values for multiple comparisons with the Bonferroni correction. The resulting significance levels are depicted in Figure 10 as coloured brackets (*p <* 0.05, *p <* 0.01, *p <* 0.001). This procedure was repeated independently for AUROC and nAUPRC, producing the four ranked panels: (A) all models / AUROC, (B) CNN-only / AUROC, (C) all models / nAUPRC, and (D) CNN-only / nAUPRC.

## Ethics

This study was conducted using three publicly available, de-identified critical care datasets: MIMIC-IV, HiRID, and eICU. The MIMIC-IV database was approved by the institutional review boards (IRBs) of the Massachusetts Institute of Technology (MIT) and Beth Israel Deaconess Medical Center (BIDMC), and is made available under a data use agreement. All users are required to complete the CITI “Data or Specimens Only Research” course before accessing the data. The dataset is de-identified in accordance with the Health Insurance Portability and Accountability Act (HIPAA). The HiRID dataset was released with approval from the Ethics Commission of the Canton of Bern (KEK), Switzerland. The data are fully de-identified and were collected retrospectively from ICU patients at the Bern University Hospital. Informed consent was waived by the ethics committee due to the anonymized nature of the data. The eICU Collaborative Research Database is a multi-center critical care database developed by Philips Healthcare in collaboration with the MIT Laboratory for Computational Physiology. It is also de-identified in compliance with HIPAA and available under a data use agreement, requiring completion of the same CITI training course for access. Since all datasets used in this study are de-identified and publicly available, and the original data collection was approved by the respective ethics committees, no additional ethics approval was required for this secondary analysis.

## Code Availability

The underlying code for this study is available in the GitLab repository “[Transfer Learning For Sepsis Prediction](https://gitlab.ethz.ch/BMDSlab/publications/sepsis/transfer-tearning-for-sepsis-detection)”.

## Data Availability

The three publicly available, de-identified datasets [HiRID](https://physionet.org/content/hirid/1.1.1/), [MIMIC-IV](https://physionet.org/content/mimiciv/3.1/) and [eICU](https://physionet.org/content/eicu-crd/2.0/) are available from [Physionet](https://physionet.org/about/database/) after successful completion of the CITI “Data or Specimens Only Research” course. Harmonized datasets can be generated using the publicly available code provided in the [YAIB repository](https://github.com/rvandewater/YAIB).

## Competing Interests

All authors declare no financial or non-financial competing interests.

## Acknowledgements

This project was supported by grant #902 of the Strategic Focus Area “Personalized Health and Related Technologies (PHRT)” of the ETH Domain and Young Investigator Grant of the Novartis Foundation for Medical-Biological Research. The funders played no role in study design, data collection, analysis and interpretation of data, or the writing of this manuscript.

ChatGPT and Claude were used for code debugging. ChatGPT and DeepL were employed for assistance with writing to improve grammar, vocabulary, and fluency.

## Author contributions statement

F.T. and L.M. conceived the experiments, L.M. and F.T. pre-processed the datasets, F.T. conducted the experiments and analysed the results. Y.F. reviewed the code base and improved it. F.T. wrote the initial version of the paper. All authors provided feedback and revised the manuscript.

## 5 Appendix

## 5.1

**Figure 12.**
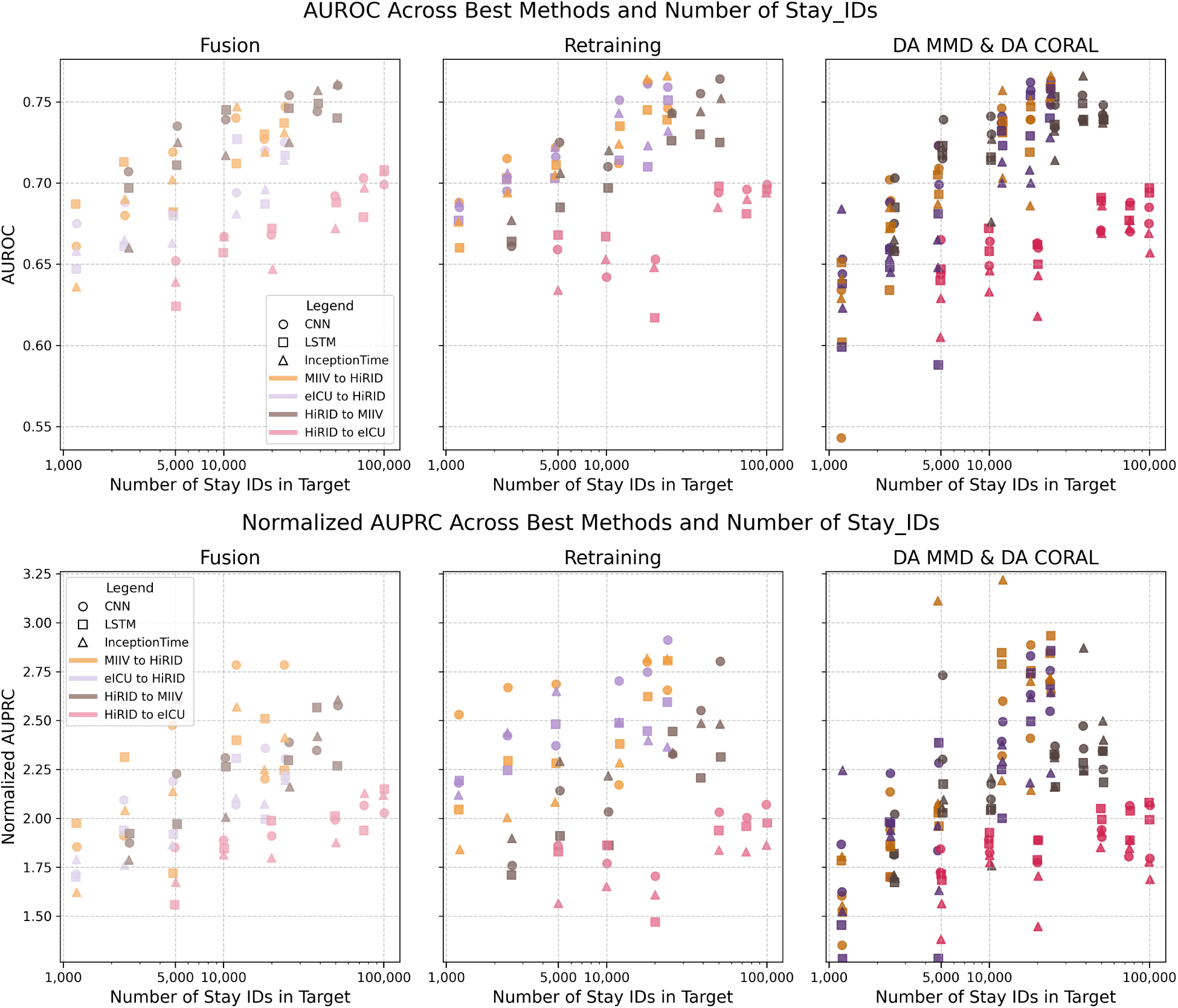
Joint DL models’ performances grouped by source-to-target combination, number of stay_ids within each training subset and transfer learning techniques.

## 5.2

**Figure 13.**
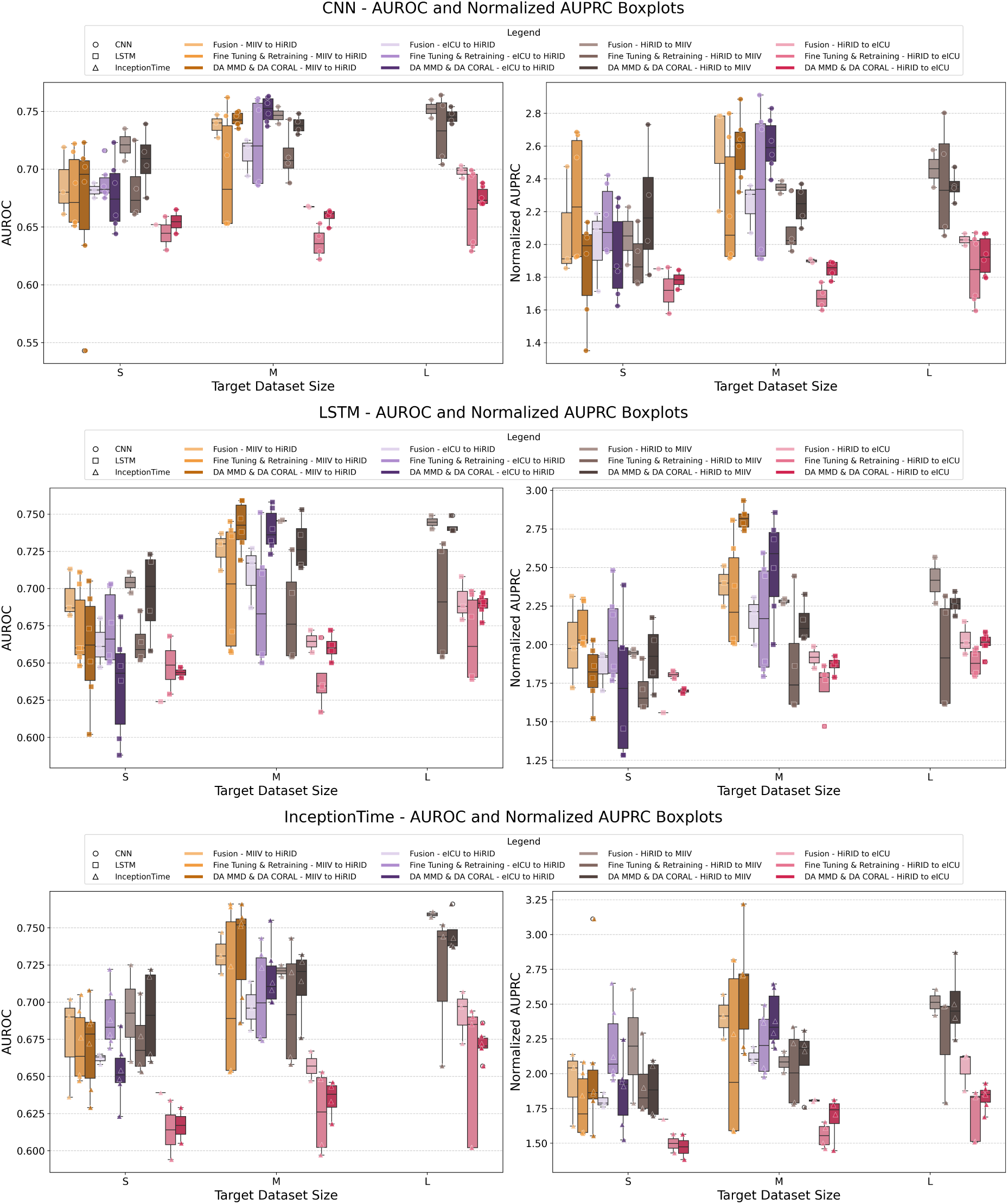
DL models’ performances grouped by source-to-target combination, target dataset size (small, medium and large) and transfer learning techniques.

**Figure 14.**
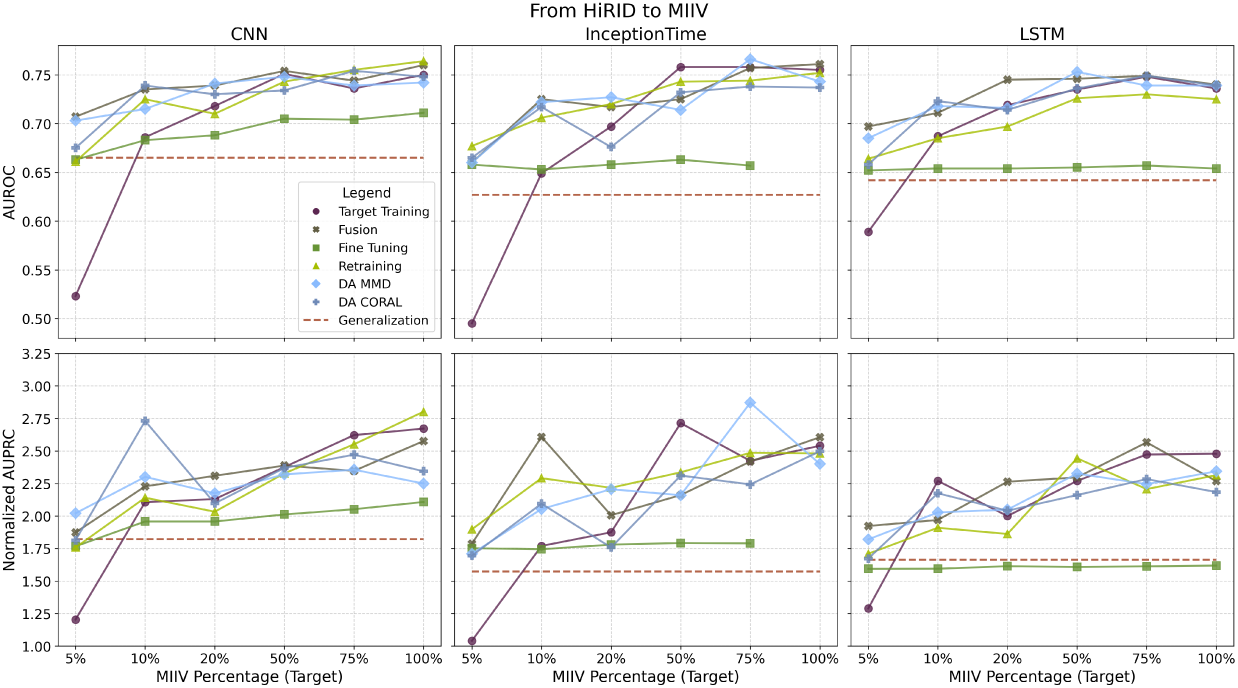
HiRID Source Dataset with MIIV Target Dataset: AUROC and nAUPRC Performances of CNN, InceptionTime and LSTM using Standard Transfer Learning, DA, Target Training and Generalization Techniques.

**Figure 15.**
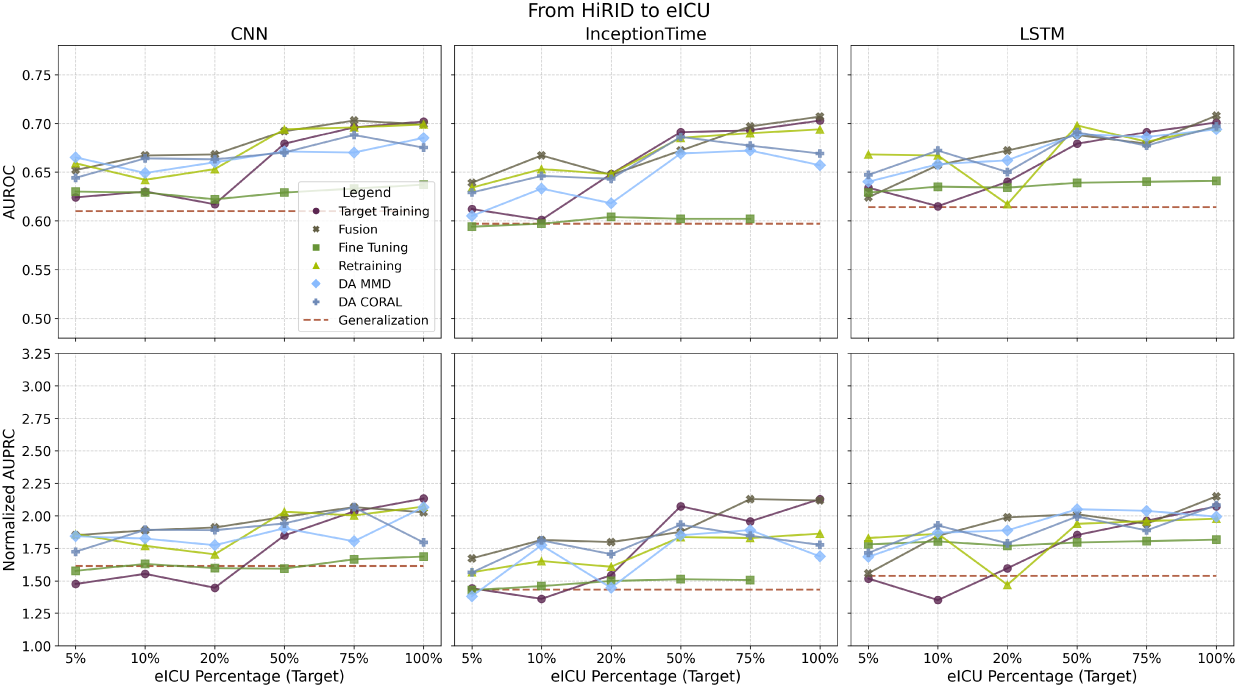
HiRID Source Dataset with eICU Target Dataset: AUROC and nAUPRC Performances of CNN, InceptionTime and LSTM using Standard Transfer Learning, DA, Target Training and Generalization Techniques.

**Figure 16.**
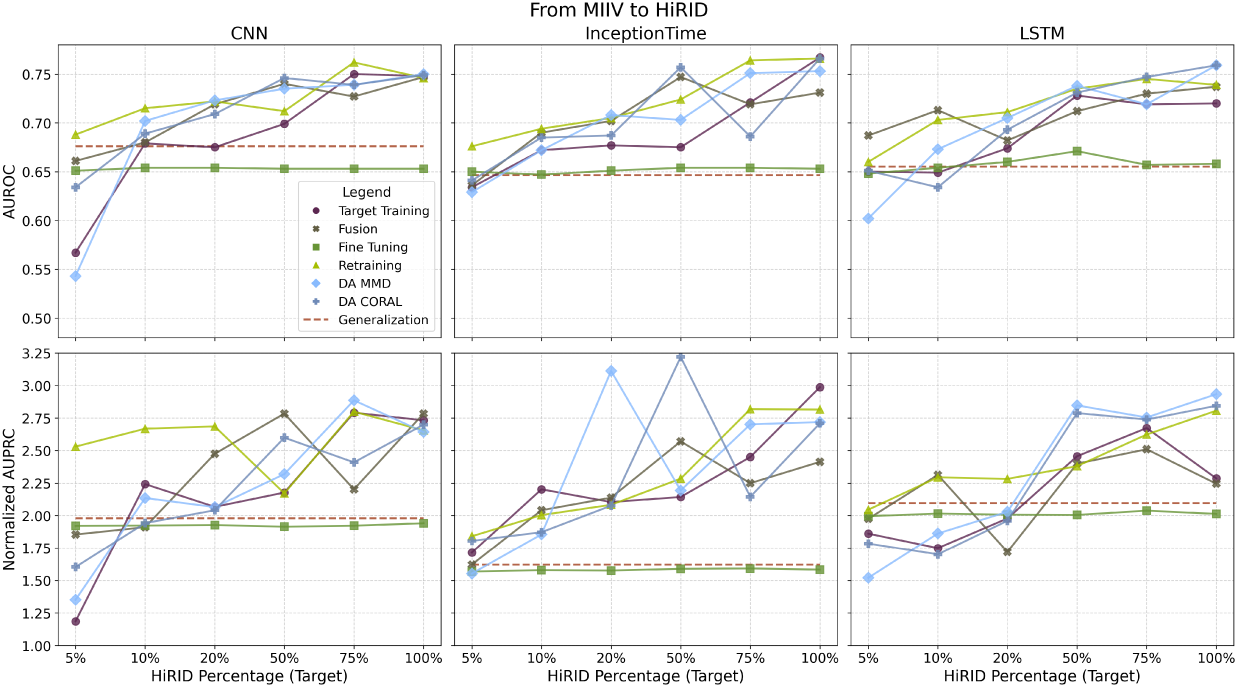
MIIV Source Dataset with HiRID Target Dataset: AUROC and nAUPRC Performances of CNN, InceptionTime and LSTM using Standard Transfer Learning, DA, Target Training and Generalization Techniques.

**Figure 17.**
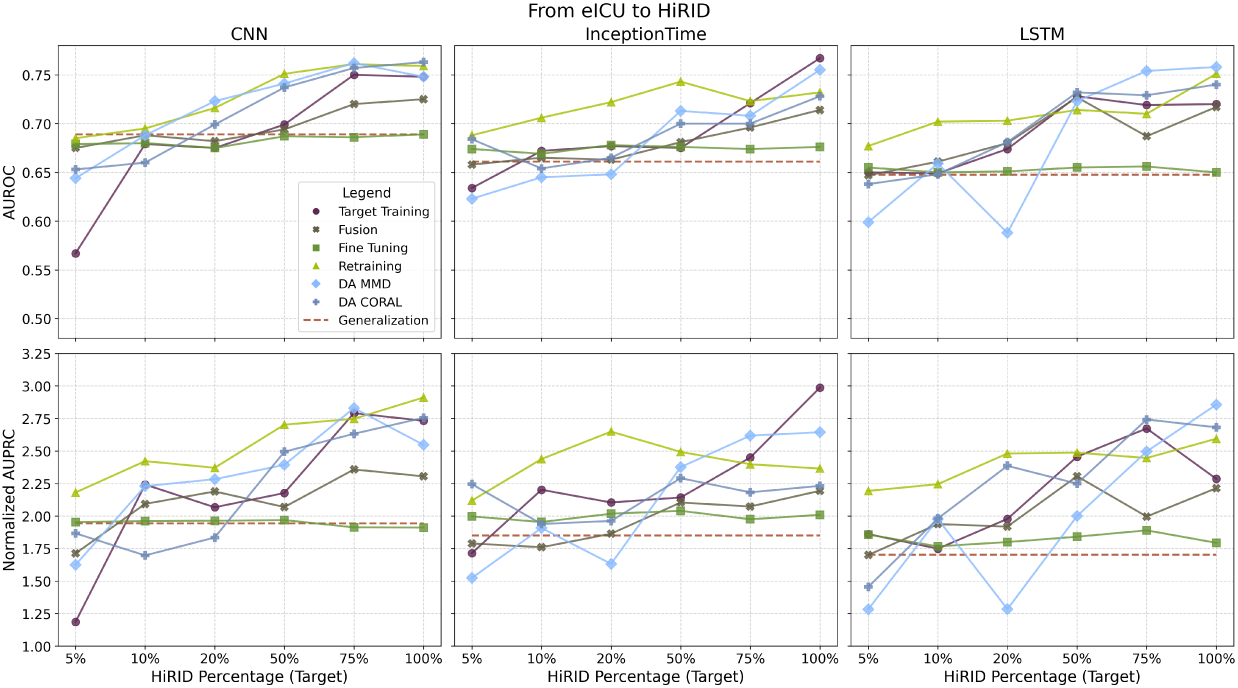
eICU Source Dataset with HiRID Target Dataset: AUROC and nAUPRC Performances of CNN, InceptionTime and LSTM using Standard Transfer Learning, DA, Target Training and Generalization Techniques.

## Notes

### Competing Interest Statement

The authors have declared no competing interest.

### Author Declarations

This study was conducted using three publicly available, de-identified critical care datasets: MIMIC-IV, HiRID, and eICU. The MIMIC-IV database was approved by the institutional review boards (IRBs) of the Massachusetts Institute of Technology (MIT) and Beth Israel Deaconess Medical Center (BIDMC), and is made available under a data use agreement. All users are required to complete the CITI Data or Specimens Only Research course before accessing the data. The dataset is de-identified in accordance with the Health Insurance Portability and Accountability Act (HIPAA). The HiRID dataset was released with approval from the Ethics Commission of the Canton of Bern (KEK), Switzerland. The data are fully de-identified and were collected retrospectively from ICU patients at the Bern University Hospital. Informed consent was waived by the ethics committee due to the anonymized nature of the data. The eICU Collaborative Research Database is a multi-center critical care database developed by Philips Healthcare in collaboration with the MIT Laboratory for Computational Physiology. It is also de-identified in compliance with HIPAA and available under a data use agreement, requiring completion of the same CITI training course for access. Since all datasets used in this study are de-identified and publicly available, and the original data collection was approved by the respective ethics committees, no additional ethics approval was required for this secondary analysis.

